# Life Satisfaction and Sleep Health among College Students in the United States

**DOI:** 10.64898/2026.09.10.26362668

**Authors:** Bethany T. Ogbenna, Wensu Zhou, LaDarius Williams, Symielle A. Gaston, Philip M. Zendels, Christopher Payne, W. Braxton Jackson, Chandra L. Jackson

## Abstract

We investigated life satisfaction in relation to sleep health among U.S. college students, and determined whether the relationship varied by age, sex, race, and ethnicity. Using 2022 National Health Interview Survey data, we assessed life satisfaction (satisfied vs. dissatisfied), recommended sleep duration (≥7 vs. <7 hours), few insomnia symptoms (IS), and restorative sleep. Adjusted Poisson regression with robust variance was employed for the overall population and then stratified by the aforementioned sociodemographic characteristics. Overall (N = 1,426, mean age ± SE = 27.7 ± 0.3) years), 97.1% of college students reported life satisfaction, which was associated with a higher prevalence of recommended sleep duration (PR = 1.96 [95% CI:1.19-3.22]), few IS (PR = 2.48 [1.36-4.52]), and restorative sleep (PR = 3.62 [1.49-8.83]). There was no evidence of differences in the relationship by age, sex, race, and ethnicity. Life satisfaction was associated with favorable sleep among U.S. college students.

## INTRODUCTION

Sleep disturbances refer to “sleep abnormalities resulting from poor sleep quality or insomnia, in the absence of other diagnosed comorbid sleep disorders”.^1, 2^ Sleep disturbances are common among students enrolled in higher education institutions worldwide.^3–7^ For instance, results from a large multi-university study of 7,626 college students conducted between 2015 and 2017 in the U.S. showed that 63% of students reported short sleep (<7 hours per night), and 43% had difficulty falling asleep (taking >30 minutes) at least once per week.^8^ Similarly, an earlier analysis of 2004-2017 U.S. National Health Interview Survey data from 2,119 college students living in dormitories reported a 29% prevalence of short sleep duration, with 19% reporting trouble falling asleep (≥3 days/week), 10% reporting trouble staying asleep (≥3 days/week), and 41% reporting non-restorative sleep (NRS, defined as waking feeling unrested 4 or more days per week).^9^ Sleep disturbances are directly associated with poorer academic performance and are also linked to symptoms of depression and anxiety, which are prevalent among college students.^10–13^ For example, according to the Healthy Minds Study in the U.S., more than 60% of college students met criteria for at least one mental health problem.^14^ Understanding the prevalence of sleep disturbances and the factors associated with them is important for informing efforts to promote student health, well-being, academic success, and flourishing.

Furthermore, sleep disturbances have been shown to differ by age, sex, and race as well as ethnicity. Traditionally, college students in the U.S. are typically aged 18-24 years and transition directly, or shortly thereafter, from high school to higher education. In contrast, non-traditional students are generally defined as people who enter higher education after the age of 24; this group may also include students in post-undergraduate education, including master’s, doctoral, and professional degree programs.^15^ Younger students, particularly first-year undergraduates (mean age = 18 years), are more likely to report insufficient sleep, later bedtimes, and irregular sleep patterns compared to their older peers (e.g., seniors, mean age=22 years).^16^ Related to age, the educational stage of college students, such as undergraduate versus post-graduate or advanced education, is an important factor that can substantially influence sleep health among college students.^17^ Irregular or early class schedules, evening coursework, heavy academic demands, and work–school conflict have been identified as factors that may adversely affect sleep among undergraduate students.^18^ Although evidence remains limited, research investigations involving master’s, doctoral, and professional graduate students such as graduate nursing students and clinical postgraduate students suggest that competing academic, work, and family responsibilities may negatively affect sleep and other health-related behaviors.^19–21^ Also, female students report more sleep disturbances than male students – partly attributable to biological (e.g., menstruation), psychological (e.g., heightened stress, rumination), and social (e.g., differential societal expectations for women pursing academic degrees) factors.^5, 16, 22–24^ Moreover, students in certain racial and ethnic groups encounter even more sleep disturbances linked to social stressors such as low socioeconomic status (SES).^9^ For instance, in a nationally representative study, Black/African American and Latino college students were more likely to report short sleep duration, IS (i.e., trouble falling and staying asleep), and NRS than Non-Hispanic (NH)-White students. NH-Asian students reported higher NRS compared to NH-White students.^9^ Identifying antecedents or factors that influence sleep health across various populations of U.S. college students is important for prevention efforts.

Life satisfaction, a key dimension of how people think and feel about their own lives, reflects overall quality of life across domains such as career, social relationships, financial circumstances, and physical health.^25–27^ It can vary across the life course and represent an important marker of psychological development, reflecting individuals’ capacity to maintain a favorable outlook on life even in the context of adversity throughout the life span.^28–32^ Similar to the variation in sleep disturbance prevalence, life satisfaction may also vary within the student population by age along with academic stage, sex, race, and ethnicity as these groups may experience different stressors and stress relievers.^33–36^ It is estimated that approximately 96% of U.S. adults aged over 18 years report life satisfaction.^37^ However, to our knowledge, no studies have estimated levels of life satisfaction among U.S. college students, assessed being satisfied with life in relation to sleep health, nor determined if the relationship differs by age, sex, race, and ethnicity.

In general vs. student populations across the globe, prior cross-sectional evidence suggests that life satisfaction and sleep are linked and may have bidirectional associations. For example, higher life satisfaction has been associated with better sleep quality and fewer sleep disturbances; conversely, poor sleep can contribute to lower life satisfaction.^38–41^ Among college students, disruptions or challenges across academic, work, family, and daily-life domains could increase stress and emotional distress, thereby reducing life satisfaction, which in turn, can be associated with sleep disturbances by prolonging sleep onset latency and fragmenting sleep.^42–44^ Some studies among university students – two conducted in Saudi Arabia and China and one involving 1,074 students at the University of North Texas in the U.S. – suggest that sufficient, high-quality sleep may enhance life satisfaction by improving emotional regulation, cognitive performance, and stress resilience.^45–47^ Evidence from relatively homogeneous samples of undergraduate students in China (n = 12,609; 68.8% freshmen, age not reported) and Israel (n = 254; mean age ± SD = 23.79 ± 1.85 years, 83% women) suggests a positive correlation between life satisfaction and better sleep quality (measured by Pittsburgh Sleep Quality Index [PSQI]).^48, 49^ However, these studies primarily relied on correlational analyses without regression modeling, limiting the robustness of conclusions.^48–50^ To our knowledge, no prior nationally representative studies of U.S. college students investigated the association between life satisfaction and sleep health.

Furthermore, it is plausible that the life satisfaction-sleep relationship differs across certain sociodemographic characteristics such as age, sex, race, and ethnicity although evidence is limited. For instance, age may reflect different developmental stages and stressors among college students, with older students potentially facing more responsibilities that may affect sleep. Additionally, there are sex differences in stress exposure, coping strategies, and emotional regulation, with women often reporting more difficulties in these domains, which may influence sex differences in relationships between life satisfaction and sleep disturbances.^16, 37^ Racial and ethnic differences in SES and social support may also shape students’ stress burden. For example, college students from different racial and ethnic groups may experience differences in financial strain, work and family responsibilities, and access to social support.^52^ These differences may, in turn, contribute to variation in both life satisfaction and sleep disturbances across racial and ethnic groups. Thus, life satisfaction may especially contribute to sleep among groups, such as older, female, and racial or ethnic minority students, who may encounter higher stress burdens and more competing demands (e.g., financial and caregiving responsibilities) or lower access to protective resources. However, the strength of associations across groups of college students is unknown. Therefore, determining whether associations between life satisfaction and sleep disturbances may differ across sociodemographic groups is important.

To address these gaps in the literature, we estimated the prevalence of life satisfaction among college students and investigated associations between life satisfaction and sleep disturbances among U.S. college students overall and by age, sex, race and ethnicity. We hypothesized that the prevalence of life satisfaction among college students is high at approximately 96%, which is comparable to prior reports among U.S. adults aged 18 years or older.^37, 53^ We further hypothesized that life satisfaction is associated with higher prevalence of recommended sleep duration, few IS, and restorative sleep and that these associations are stronger among older versus younger college students, women versus men, and Latino and NH-Black/African American versus NH-White students. As an exploratory aim, we additionally stratified by age, sex, and student status within each racial and ethnic group to assess effect modification within racial and ethnic groups. As a sensitivity analysis, we further examined overall and race along with ethnicity-stratified associations by student status.

## METHODS

### Data source: The National Health Interview Survey

The data were accessed through the Integrated Public Use Microdata Series.^54^ We used cross-sectional data from the 2022 National Health Interview Survey (NHIS), a nationally representative survey of the non-institutionalized U.S. population aged 18 years of age and older residing in the 50 states and the District of Columbia. The 2022 survey year was selected because it represented the earliest and only available year for which data on life satisfaction and sleep were simultaneously available at the time of analysis. The NHIS employed a complex, multistage probability sampling design that included stratification and clustering. Data on sociodemographic and health characteristics were collected through face-to-face interviews. Detailed descriptions of the methodology are available elsewhere.^55^ For 2022 NIHS, the final sample adult response rate was 47.7%.^56, 57^ Informed consent was obtained from all participants, and approval for the current study of these publicly available, de-identified data was waived by the Institutional Review Board of the National Institutes of Health since it is not considered human subjects research.

### Study population

The study population included 1,426 college students, defined as adults who reported completing high school or more and currently attending school. Supplemental Figure 1 outlines the sample selection process. Among otherwise eligible college students, excluded participants were few (n = 32). The main reasons for exclusion were missing data on sleep measures, life satisfaction, or other covariates. Compared with participants included in the analytic sample, excluded participants were younger (24.0 [SE=1.7] years vs. 27.7 [SE=0.3] years), more likely to be undergraduate students (based on educational attainment of <college), and had a higher prevalence of recommended sleep duration and few IS (Supplemental Table 1). There were no other significant differences between included and excluded participants.

### Exposure assessment: Life satisfaction

Satisfaction with life was defined using the question: “In general, how satisfied are you with your life? Would you say very satisfied, satisfied, dissatisfied, or very dissatisfied?”. Participants self-reported being ‘very satisfied’ or ‘satisfied’ were classified as having satisfaction with life, while participants who reported ‘dissatisfied’ or ‘very dissatisfied’ were classified as having dissatisfaction with life.

### Outcome assessment: Sleep health

Sleep duration was measured using the question: “On average, how many hours of sleep do you get in a 24-hour period?” Responses were recorded as whole numbers rounded to the nearest hour. Sleep duration was categorized as recommended versus not recommended (≥7 hours vs. <7 hours), following the guidelines from the American Academy of Sleep Medicine and Sleep Research Society.^58^ IS were measured using two questions: 1) “During the past 30 days, how often did you have trouble falling asleep?” and 2) “During the past 30 days, how often did you have trouble staying asleep?” Response options for each item were “never,” “some days,” “most days,” and “every day.”^59^ Few IS was defined as having trouble falling and staying asleep “never/some days” versus “most days/every day” in the past 30 days.^60^ Restorative sleep was assessed using the question: “During the past 30 days, how often did you wake up feeling well-rested?”. Restorative sleep was defined as feeling rested upon waking versus “most days/every day” versus “never/some days” in the past 30 days.^60^

### Potential confounders

Based on prior literature and construction of a directed acyclic graph (Supplemental Figure 2),^61^ the following variables were included as potential confounders: age (years; when not stratified); sex (men, women) when not stratified; race and ethnicity (Hispanic/Latino [hereafter Latino], NH-Black/African American, NH-Other [including Asian, American Indian/Alaska Native, multiracial, and other race and ethnicity [hereafter, NH-Other including Asian, American Indian/Alaska Native, multiracial, and other races]), and NH-White when not stratified; student status (undergraduate student [defined as highest educational attainment <college] versus graduate student [educational attainment ≥college]) when not stratified; employment status (employed, not employed); ratio of family income to the poverty threshold (<1.00, 1.00–1.99, 2.00–2.99, ≥3.00); marital status (divorced/widowed/separated/married–spouse absent, single/never married, married–spouse present/living with partner); region of residence (Northeast, Midwest, South, West); health behaviors, including leisure-time physical activity (inactive, insufficiently active, sufficiently active) defined using the 2018 U.S. Department of Health and Human Services Physical Activity guidelines;^62^ smoking status (never/quit > 12 months prior to interview, former, current); alcohol consumption (current, former, lifetime abstainer); body mass index (BMI) (underweight (<18.5 kg/m^2^), recommended (18.5-24.9 kg/m^2^), overweight (25-29.9 kg/m^2^), obesity (≥30 kg/m^2^)); depression (yes, no); and general health status (excellent/very good/good, fair/poor). Depression was assessed based on whether respondents reported having ever been told by a doctor or other health professional that they had persistent depressive disorder, postpartum depression, psychotic depression, seasonal affective disorder, or bipolar disorder.

### Potential effect modifiers

Potential effect modifiers included age category (<23 years, ≥23 years), sex (men, women), race and ethnicity (Latino, NH-Black/African American, NH-Other, and NH-White), and student status (undergraduate, graduate).

### Statistical analysis

Descriptive statistics for the study population were estimated overall, by satisfaction with life (satisfaction vs. dissatisfaction), and by satisfaction with life within racial and ethnic groups. We reported sample weighted means (standard errors [SEs]) for age and age-standardized (based on the 2010 U.S. Census population), sample weighted proportions for categorical variables. The NHIS sampling weights used in these analyses account for the inverse probability of selection to reflect the complex survey design. We also reported descriptive statistics stratified by age category, sex, and student status. Further, we assessed the distribution of life satisfaction by age category, sex, race and ethnicity. These descriptive analyses show any variation in life satisfaction across sociodemographic group and support the interpretation of effect modification.

Poisson regression with robust variance was used to estimate prevalence ratios (PRs) and 95% confidence intervals (CIs) for associations between life satisfaction and sleep disturbances adjusting for potential confounders in the overall models. Model 1 adjusted for age (years), sex, student status, employment status, ratio of family income to poverty threshold, marital status, region of residence, and general health status. Model 2, additionally adjusted for smoking status, alcohol consumption, leisure-time physical activity, BMI, and depression. Models were stratified by potential effect modifiers: age category, sex, and race along with ethnicity. To test effect modification by age, sex, and race as well as ethnicity, separately, cross-product terms (i.e., life satisfaction*age, life satisfaction*sex, and life satisfaction*race and ethnicity) were included in the overall model. Cross-product terms (i.e., life satisfaction*age, life satisfaction*sex) were also added to the models within racial and ethnic groups to test for effect modification within racial and ethnic groups. Lastly, using the same approach to testing effect modification, we further examined whether associations varied by student status as a sensitivity analysis. The sensitivity analysis was used to determine whether results are robust across undergraduate and graduate students. All analyses were performed using estimation and post-estimation commands for survey data in Stata, Version 15.1 (Statacorp, College Station, Texas), and a two-sided p-value of 0.05 was used to determine statistical significance.

## RESULTS

### Study population characteristics

Table 1 lists sociodemographic, health behavior, and clinical characteristics among college students, overall and by race and ethnicity. Among a total of 1,426 participants, the mean age was 27.7 years (SE = 0.31) with less participants aged <23 years (45.8%) vs. ≥23 years (54.2%) and more women than men (61.4% vs. 38.6%). By race and ethnicity, participants identified as Hispanic/Latino (21.0%); NH-Black/African American (14.0%); NH and another race (14.0%, including non-Hispanic, Asian, American Indian/Alaska Native, multiracial, and other races); or NH-White (51.0%). Data for the heterogenous NH and another race group are provided but not interpreted due to heterogeneity in the group. Based on reports of prior educational attainment, a higher proportion of participants were undergraduate students (58.5%) than graduate students (41.5%).

**Table 1.** Age-standardized sociodemographic, health behavior, and clinical characteristics among college students, overall and by race and ethnicity, National Health Interview Survey, 2022, (N=1,426)

|  | Total<br>n=1,426 (100%) |  |  |  | Hispanic/Latine<br>n=299 (21.0%) |  |  |  | NH-Black/<br>African American<br>n=200 (14.0%) |  |  |  | NH-Other<br>n=200 (14.0%) |  |  |  | NH-White<br>n=727 (51.0%) |  |  |  |
| --- | --- | --- | --- | --- | --- | --- | --- | --- | --- | --- | --- | --- | --- | --- | --- | --- | --- | --- | --- | --- |
|  | All<br>n=1,426<br>(100%) | Life satisfaction <sup>a</sup><br>Yes<br>n=1,385<br>(97.1%) | No<br>n=41<br>(2.9%) | Chi-square<br>or t-test<br>p-value | All<br>n=299<br>(100%) | Life satisfaction <sup>a</sup><br>Yes<br>n=288<br>(96.3%) | No<br>n=11<br>(3.7%) | Chi-square<br>or t-test<br>p-value | All<br>n=200<br>(100%) | Life satisfaction <sup>a</sup><br>Yes<br>n=195<br>(97.5%) | No<br>n=5<br>(2.5%) | Chi-square<br>or t-test<br>p-value | All<br>n=200<br>(100%) | Life satisfaction <sup>a</sup><br>Yes<br>n=197<br>(98.5%) | No<br>n=3<br>(1.5%) | Chi-square<br>or t-test<br>p-value | All<br>n=727<br>(100%) | Life satisfaction <sup>a</sup><br>Yes<br>n=705<br>(97.0%) | No<br>n=22<br>(3.0%) | Chi-square<br>or t-test<br>p-value |
| **Sociodemographic Characteristics** |  |  |  |  |  |  |  |  |  |  |  |  |  |  |  |  |  |  |  |  |
| Age (years), mean (SE) | 27.7<br>(0.3) | 27.7<br>(0.3) | 26.9<br>(1.3) | 0.566 | 25.3<br>(0.5) | 25.2<br>(0.5) | 26.4<br>(2.7) | 0.682 | 29.1<br>(0.8) | 29.4<br>(0.8) | 21.8<br>(1.6) | <0.001 | 26.2<br>(0.7) | 26.2<br>(0.7) | 30.1<br>(3.2) | 0.230 | 28.6<br>(0.5) | 28.6<br>(0.5) | 28.4<br>(2.0) | 0.923 |
| Age group |  |  |  | 0.580 |  |  |  | 0.826 |  |  |  | 0.418 |  |  |  | 0.088 |  |  |  | 0.393 |
| <23 years | 45.8 | 46.0 | 40.5 |  | 50.4 | 50.3 | 54.3 |  | 34.3 | 33.7 | 53.4 |  | 51.9 | 52.6 | 0.0 |  | 45.6 | 45.9 | 34.1 |  |
| ≥23 years | 54.2 | 54.0 | 59.5 |  | 49.6 | 49.7 | 45.7 |  | 65.7 | 66.3 | 46.6 |  | 48.1 | 47.4 | 100.0 |  | 54.4 | 54.1 | 65.9 |  |
| Sex |  |  |  | 0.136 |  |  |  | 0.298 |  |  |  | 0.058 |  |  |  | 0.924 |  |  |  | 0.521 |
| Men | 38.6 | 38.4 | 50.2 |  | 29.0 | 28.6 | 37.6 |  | 33.9 | 33.3 | 73.1 |  | 29.2 | 29.3 | 31.9 |  | 43.1 | 42.9 | 50.7 |  |
| Women | 61.4 | 61.6 | 49.8 |  | 71.0 | 71.4 | 62.4 |  | 66.1 | 66.7 | 26.9 |  | 70.8 | 70.7 | 68.1 |  | 56.9 | 57.1 | 49.3 |  |
| Student status |  |  |  | 0.273 |  |  |  | 0.340 |  |  |  | 0.031 |  |  |  | 0.487 |  |  |  | 0.265 |
| Undergraduate students | 58.5 | 58.5 | 65.5 |  | 81.4 | 81.9 | 51.9 |  | 57.8 | 57.5 | 91.9 |  | 73.2 | 73.3 | 53.7 |  | 52.6 | 52.4 | 66.3 |  |
| Graduate students | 41.5 | 41.5 | 34.5 |  | 18.6 | 18.1 | 48.1 |  | 42.2 | 42.5 | 8.1 |  | 26.8 | 26.7 | 46.3 |  | 47.4 | 47.6 | 33.7 |  |
| Employment status |  |  |  | 0.176 |  |  |  | 0.249 |  |  |  | 0.509 |  |  |  | 0.100 |  |  |  | 0.361 |
| Employed | 59.5 | 60.0 | 44.7 |  | 70.3 | 70.9 | 40.4 |  | 62.8 | 63.2 | 46.6 |  | 54.4 | 54.7 | 0.0 |  | 59.3 | 59.8 | 48.2 |  |
| Not employed | 40.5 | 40.0 | 55.3 |  | 29.7 | 29.1 | 59.6 |  | 37.2 | 36.8 | 53.4 |  | 45.6 | 45.3 | 100.0 |  | 40.7 | 40.2 | 51.8 |  |
| Ratio of family income to poverty threshold |  |  |  | 0.001 |  |  |  | 0.649 |  |  |  | 0.177 |  |  |  | 0.471 |  |  |  | <0.001 |
| <1.00 | 7.8 | 7.6 | 22.6 |  | 17.2 | 17.3 | 9.0 |  | 8.2 | 8.4 | 0.0 |  | 11.9 | 11.6 | 68.1 |  | 6.0 | 5.6 | 27.6 |  |
| 1.00-1.99 | 15.1 | 14.9 | 35.7 |  | 25.1 | 24.7 | 53.4 |  | 20.3 | 20.6 | 8.1 |  | 15.5 | 15.6 | 0.0 |  | 12.6 | 12.2 | 35.4 |  |
| 2.00-2.99 | 16.0 | 15.9 | 15.5 |  | 18.7 | 18.6 | 19.5 |  | 23.5 | 22.7 | 65.0 |  | 25.8 | 25.8 | 31.9 |  | 11.5 | 11.7 | 5.3 |  |
| ≥3.00 | 61.1 | 61.6 | 26.2 |  | 38.9 | 39.4 | 18.1 |  | 48.0 | 48.3 | 26.9 |  | 46.8 | 47.0 | 0.0 |  | 69.9 | 70.6 | 31.7 |  |
| Marital status |  |  |  | 0.002 |  |  |  | 0.062 |  |  |  | 0.474 |  |  |  | 0.508 |  |  |  | 0.060 |
| Divorced/Widowed/Separated/<br>Married-spouse absent | 18.7 | 18.5 | 21.6 |  | 9.5 | 9.7 | 0.0 |  | 24.3 | 24.4 | 0.0 |  | 25.9 | 25.9 | 0.0 |  | 16.8 | 16.5 | 27.0 |  |
| Single/Never married | 36.9 | 36.5 | 66.0 |  | 47.4 | 46.4 | 94.3 |  | 45.0 | 44.6 | 100.0 |  | 45.4 | 45.4 | 78.2 |  | 33.5 | 33.3 | 53.0 |  |
| Married-spouse<br>present/Living with partner | 44.4 | 44.9 | 12.4 |  | 43.1 | 43.9 | 5.7 |  | 30.6 | 31.0 | 0.0 |  | 28.7 | 28.7 | 21.8 |  | 49.6 | 50.2 | 20.1 |  |
| Region of residence |  |  |  | 0.715 |  |  |  | 0.074 |  |  |  | 0.581 |  |  |  | 0.593 |  |  |  | 0.737 |
| Northeast | 16.2 | 16.1 | 21.2 |  | 19.8 | 20.0 | 6.2 |  | 14.8 | 14.5 | 37.9 |  | 12.4 | 12.5 | 0.0 |  | 16.7 | 16.5 | 24.8 |  |
| Midwest | 22.0 | 22.0 | 24.2 |  | 5.4 | 5.6 | 0.0 |  | 20.1 | 19.9 | 26.9 |  | 19.6 | 19.8 | 0.0 |  | 25.8 | 25.7 | 30.3 |  |
| South | 34.7 | 35.0 | 24.0 |  | 32.1 | 32.7 | 8.6 |  | 52.1 | 52.4 | 35.2 |  | 30.4 | 30.1 | 78.2 |  | 33.7 | 34.0 | 25.1 |  |
| West | 27.1 | 26.9 | 30.6 |  | 42.7 | 41.6 | 85.2 |  | 13.1 | 13.3 | 0.0 |  | 37.5 | 37.6 | 21.8 |  | 23.9 | 23.8 | 19.8 |  |
| **Health Behaviors** |  |  |  |  |  |  |  |  |  |  |  |  |  |  |  |  |  |  |  |  |
| Smoking status |  |  |  | 0.184 |  |  |  | 0.921 |  |  |  | 0.773 |  |  |  | 0.977 |  |  |  | 0.037 |
| Never/quit>12 months prior | 95.0 | 95.2 | 85.9 |  | 94.3 | 94.4 | 94.0 |  | 89.0 | 88.9 | 100.0 |  | 98.3 | 98.3 | 100.0 |  | 95.8 | 96.0 | 82.9 |  |
| Former/quit≤12 months ago | 0.8 | 0.8 | 2.3 |  | 0.6 | 0.6 | 0.0 |  | 0.6 | 0.6 | 0.0 |  | 0.6 | 0.6 | 0.0 |  | 0.9 | 0.8 | 4.7 |  |
| Current | 4.2 | 4.1 | 11.8 |  | 5.1 | 5.0 | 6.0 |  | 10.5 | 10.5 | 0.0 |  | 1.1 | 1.1 | 0.0 |  | 3.4 | 3.2 | 12.5 |  |
| Alcohol consumption |  |  |  | 0.445 |  |  |  | 0.600 |  |  |  | 0.239 |  |  |  | 0.453 |  |  |  | 0.910 |
| Current (≥1 drink past year) | 67.0 | 66.8 | 76.3 |  | 67.6 | 67.2 | 84.6 |  | 55.7 | 55.1 | 100.0 |  | 40.3 | 40.4 | 46.3 |  | 73.1 | 73.1 | 76.2 |  |
| Former (no drinks past year) | 17.2 | 17.2 | 10.3 |  | 10.8 | 11.1 | 0.0 |  | 28.4 | 28.5 | 0.0 |  | 26.1 | 25.9 | 53.7 |  | 14.9 | 15.0 | 11.2 |  |
|  | All<br>n=1,426<br>(100%) | Life satisfaction <sup>a</sup><br>Yes<br>n=1,385<br>(97.1%) | No<br>n=41<br>(2.9%) | Chi-square<br>or t-test<br>p-value | All<br>n=299<br>(100%) | Life satisfaction <sup>a</sup><br>Yes<br>n=288<br>(96.3%) | No<br>n=11<br>(3.7%) | Chi-square<br>or t-test<br>p-value | All<br>n=200<br>(100%) | Life satisfaction <sup>a</sup><br>Yes<br>n=195<br>(97.5%) | No<br>n=5<br>(2.5%) | Chi-square<br>or t-test<br>p-value | All<br>n=200<br>(100%) | Life satisfaction <sup>a</sup><br>Yes<br>n=197<br>(98.5%) | No<br>n=3<br>(1.5%) | Chi-square<br>or t-test<br>p-value | All<br>n=727<br>(100%) | Life satisfaction <sup>a</sup><br>Yes<br>n=705<br>(97.0%) | No<br>n=22<br>(3.0%) | Chi-square or<br>t-test<br>p-value |
| Lifetime abstinence (<12 drinks in life) | 15.8 | 15.9 | 13.4 |  | 21.6 | 21.7 | 15.4 |  | 15.9 | 16.4 | 0.0 |  | 33.6 | 33.8 | 0.0 |  | 12.0 | 12.0 | 12.7 |  |
| Leisure-time physical activity |  |  |  | 0.040 |  |  |  | <0.001 |  |  |  | 0.437 |  |  |  | 0.636 |  |  |  | 0.142 |
| Inactive | 18.0 | 17.6 | 41.0 |  | 13.2 | 11.8 | 74.0 |  | 22.2 | 22.4 | 0.0 |  | 22.0 | 22.1 | 0.0 |  | 16.4 | 16.0 | 33.7 |  |
| Insufficiently active | 25.2 | 25.2 | 24.2 |  | 31.3 | 32.0 | 3.2 |  | 22.3 | 22.4 | 15.5 |  | 47.3 | 46.8 | 100.0 |  | 21.7 | 21.7 | 26.0 |  |
| Sufficiently active | 56.8 | 57.2 | 34.8 |  | 55.4 | 56.2 | 22.8 |  | 55.6 | 55.2 | 84.5 |  | 30.7 | 31.0 | 0.0 |  | 61.9 | 62.3 | 40.3 |  |
| Usual sleep duration |  |  |  | 0.009 |  |  |  | 0.220 |  |  |  | 0.782 |  |  |  | 0.023 |  |  |  | 0.045 |
| Short (<7 hours) | 29.3 | 28.9 | 47.1 |  | 35.2 | 34.5 | 46.0 |  | 54.5 | 54.2 | 62.1 |  | 30.6 | 30.1 | 100.0 |  | 24.1 | 23.7 | 44.2 |  |
| Recommended (≥7 hours) | 70.7 | 71.1 | 52.9 |  | 64.8 | 65.5 | 54.0 |  | 45.5 | 45.8 | 37.9 |  | 69.4 | 69.9 | 0.0 |  | 75.9 | 76.3 | 55.8 |  |
| Few insomnia (yes) <sup>b</sup> | 76.6 | 77.3 | 37.4 | <0.001 | 72.0 | 73.8 | 6.2 | <0.001 | 85.5 | 86.0 | 46.0 | 0.123 | 78.5 | 78.9 | 21.8 | 0.012 | 75.3 | 76.0 | 42.2 | 0.008 |
| Restorative sleep (yes) <sup>c</sup> | 61.0 | 61.9 | 8.0 | <0.001 | 54.4 | 55.6 | 6.3 | <0.001 | 60.4 | 60.6 | 35.2 | 0.296 | 58.6 | 59.0 | 0.0 | <0.001 | 61.7 | 62.8 | 5.5 | <0.001 |
| **Clinical Characteristics** |  |  |  |  |  |  |  |  |  |  |  |  |  |  |  |  |  |  |  |  |
| Ever had depression (yes) <sup>d</sup> | 23.2 | 22.4 | 68.5 | <0.001 | 14.0 | 12.9 | 65.5 | <0.001 | 11.3 | 11.1 | 26.9 | 0.406 | 21.3 | 21.0 | 78.2 | 0.012 | 27.9 | 27.0 | 71.6 | <0.001 |
| Body mass index category |  |  |  | 0.015 |  |  |  | <0.001 |  |  |  | 0.270 |  |  |  | 0.614 |  |  |  | 0.033 |
| Underweight (<18.5 kg/m<sup>2</sup>) | 1.5 | 1.4 | 9.6 |  | 1.4 | 0.7 | 41.6 |  | 2.0 | 2.1 | 0.0 |  | 4.7 | 4.7 | 0.0 |  | 1.2 | 1.1 | 8.3 |  |
| Recommended (18.5-<25 kg/m<sup>2</sup>) | 36.8 | 37.0 | 24.7 |  | 32.2 | 32.2 | 28.7 |  | 31.1 | 30.4 | 76.3 |  | 54.6 | 54.9 | 0.0 |  | 36.7 | 37.2 | 18.7 |  |
| Overweight (25-<30 kg/m<sup>2</sup>) | 32.7 | 32.6 | 39.5 |  | 40.6 | 41.3 | 9.4 |  | 28.9 | 28.9 | 23.7 |  | 23.8 | 23.6 | 53.7 |  | 32.4 | 32.1 | 45.8 |  |
| Obese (≥30 kg/m<sup>2</sup>) | 28.9 | 29.0 | 26.2 |  | 25.8 | 25.9 | 20.3 |  | 38.0 | 38.6 | 0.0 |  | 17.0 | 16.8 | 46.3 |  | 29.6 | 29.7 | 27.2 |  |
| General health status |  |  |  | <0.001 |  |  |  | 0.423 |  |  |  | 0.339 |  |  |  | <0.001 |  |  |  | 0.001 |
| Excellent/Very good/Good | 93.1 | 93.5 | 71.3 |  | 94.2 | 94.4 | 90.7 |  | 96.0 | 96.2 | 88.7 |  | 96.8 | 97.2 | 21.8 |  | 91.9 | 92.5 | 65.9 |  |
| Fair/poor | 6.9 | 6.5 | 28.7 |  | 5.8 | 5.6 | 9.3 |  | 4.0 | 3.8 | 11.3 |  | 3.2 | 2.8 | 78.2 |  | 8.1 | 7.5 | 34.1 |  |

Abbreviations: SE (standard error), NH (non-Hispanic), PA (physical activity)
Note: All estimates are age-standardized to the US 2020 population, except for age and weighted for the survey's complex sampling design. Data are presented as column percentages or means and standard errors. Percentages may not sum to 100 due to rounding or missing data. College students are defined as having completed High School or more (EDUC) and currently attending school (SCHOOLNOW)
<sup>a</sup> Life satisfaction defined as a response of 'very dissatisfied' or 'dissatisfied' vs. 'satisfied' or 'very satisfied'. 'In general, how satisfied are you with your life? Are you very satisfied, satisfied, dissatisfied, or very dissatisfied?'
<sup>b</sup> Few insomnia defined as not having trouble falling and staying asleep 'most days' or 'every day' in the past 30 days
<sup>c</sup> Restorative sleep defined as feeling rested upon waking 'most days' or 'every day' in the past 30 days
<sup>d</sup> Depression was defined by the 2019 Field Representative's Manual defines depression (major depressive disorder or clinical depression) as a common but serious mood disorder. It causes severe symptoms that affect how you feel, think, and handle daily activities, such as sleeping, eating, or working.

Life satisfaction was reported by 97.1% of participants. Compared to participants who did not report life satisfaction, participants who reported life satisfaction were more likely to have a ratio of family income to poverty ratio ≥3.00 (61.6% vs. 26.2%), to be married with a spouse present or living with a partner (44.9% vs. 12.4%), and to report sufficient leisure-time physical activity (57.2% vs. 34.8%). The unadjusted prevalence of life satisfaction did not vary significantly by age, sex, or race and ethnicity (all p-values for group comparisons >0.05, Table 1 and Supplemental Tables 2-3). Within Latino, NH-Black/African American, and NH-White groups, the prevalence of life satisfaction did not differ significantly by age or sex. Detailed age-standardized distributions of sociodemographic, health behavior, and clinical characteristics among college students, overall and stratified by age and sex are presented in Supplemental Tables 4-5.

Overall, the prevalence of recommended sleep duration, few IS, and restorative sleep was 70.7%, 76.6%, and 61.0%, respectively (Table 1). Participants who reported life satisfaction versus dissatisfaction had a higher prevalence of reporting recommended sleep duration (71.1%vs. 52.9%), few IS (77.3% vs. 37.4%), and restorative sleep (61.9% vs. 8.0%).

### Overall associations between life satisfaction and sleep

Life satisfaction was associated with an 96% higher prevalence of recommended sleep duration (aPR = 1.96, 95% CI: 1.19-3.22), an approximately 2.5-fold higher prevalence of few IS (aPR = 2.48, 95% CI: 1.36-4.52), and above a 3.5-fold higher prevalence of restorative sleep (aPR = 3.62, 95% CI: 1.49-8.33) after adjusting for age, sex, race and ethnicity, student status, employment status, ratio of family income to poverty threshold, marital status, region of residence, smoking status, alcohol consumption, leisure-time physical activity, depression, BMI, and general health status (Tables 2-3; Model 2).

**Table 2.** Prevalence ratios for associations between life satisfaction and sleep among college students, overall and stratified by race and ethnicity and age, National Health Interview Survey, 2022, (N=1,426)

|  | Recommended sleep duration<br>(≥7 hours) vs. Short (<7 hours) |  | Few insomnia symptoms<br>(yes vs. no)<sup>a</sup> |  | Restorative sleep<br>(yes vs. no)<sup>b</sup> |  |
| --- | --- | --- | --- | --- | --- | --- |
|  | Model 1<sup>c</sup> | Model 2<sup>c</sup> | Model 1<sup>c</sup> | Model 2<sup>c</sup> | Model 1<sup>c</sup> | Model 2<sup>c</sup> |
|  | Prevalence Ratio (95% Confidence Interval) for Associations with Life satisfaction<sup>d</sup> (satisfied vs. dissatisfied) |  |  |  |  |  |
| **Overall (n=1,426)** | **2.09**<br>**(1.28 - 3.42)** | **1.96**<br>**(1.19 - 3.22)** | **2.77**<br>**(1.51 - 5.08)** | **2.48**<br>**(1.36 - 4.52)** | **4.43**<br>**(1.77 - 11.1)** | **3.62**<br>**(1.49 - 8.83)** |
| <23 years (n=457) | 2.98<br>(0.89 - 9.95) | 2.78<br>(0.87 - 8.87) | 2.73<br>**(1.03 - 7.22)** | 2.53<br>(0.99 - 6.41) | 3.80<br>(0.91 - 15.9) | 3.31<br>(0.85 - 12.9) |
| ≥23 years (n=969) | **1.75**<br>**(1.09 - 2.83)** | 1.64<br>(0.99 - 2.69) | **2.64**<br>**(1.38 - 5.06)** | **2.36**<br>**(1.18 - 4.70)** | **4.53**<br>**(1.33 - 15.4)** | **3.69**<br>**(1.15 - 11.9)** |
| **Hispanic/Latine (n=299)** | **3.21**<br>**(1.12 - 9.21)** | 3.07<br>(0.96 - 9.86) | **7.89**<br>**(1.86 - 33.5)** | **7.29**<br>**(1.61 - 33.1)** | **5.58**<br>**(1.36 - 22.9)** | **4.86**<br>**(1.14 - 20.7)** |
| <23 years (n=116) | NE | NE | 8.46<br>**(1.05 - 68.4)** | 8.65<br>**(1.01 - 73.8)** | 4.72<br>(0.57 - 39.5) | 4.44<br>(0.50 - 39.3) |
| ≥23 years (n=183) | 1.42<br>(0.60 - 3.35) | 1.33<br>(0.44 - 4.06) | 7.14<br>(0.95 - 53.5) | 6.57<br>(0.76 - 56.7) | 5.88<br>(0.84 - 41.0) | 5.46<br>(0.58 - 51.7) |
| **NH-Black/African American (n=200)** | 1.67<br>(0.43 - 6.45) | 1.93<br>(0.44 - 8.34) | 1.75<br>(0.63 - 4.84) | 1.76<br>(0.66 - 4.68) | 1.25<br>(0.37 - 4.22) | 1.17<br>(0.33 - 4.17) |
| <23 years (n=47) | 1.30<br>(0.70 - 2.42) | 1.41<br>(0.58 - 3.44) | 1.37<br>(0.70 - 2.70) | 1.56<br>(0.84 - 2.88) | NE | NE |
| ≥23 years (n=153) | NE | NE | 3.80<br>(0.58 - 25.1) | 4.03<br>(0.56 - 29.1) | 0.70<br>(0.28 - 1.73) | 0.56<br>(0.24 - 1.32) |
| **NH-Other (n=200)** | NE | NE | 4.34<br>(0.61 - 30.7) | 3.61<br>(0.56 - 23.4) | NE | NE |
| <23 years (n=80) | NE | NE | NE | NE | NE | NE |
| ≥23 years (n=120) | NE | NE | 4.41<br>(0.61 - 31.6) | 2.08<br>(0.39 - 11.2) | NE | NE |
| **NH-White (n=727)** | **1.72**<br>**(1.03 - 2.87)** | 1.63<br>(0.98 - 2.72) | **2.32**<br>**(1.08 - 4.96)** | 2.01<br>(0.92 - 4.40) | **6.74**<br>**(1.16 - 39.3)** | 5.25<br>(0.97 - 28.5) |
| <23 years (n=214) | 3.09<br>(0.53 - 18.0) | 2.97<br>(0.66 - 13.4) | 2.91<br>(0.54 - 15.5) | 2.78<br>(0.60 - 12.9) | 2.36<br>(0.49 - 11.46) | 2.05<br>(0.55 - 7.67) |
| ≥23 years (n=513) | 1.40 | 1.35 | 1.93 | 1.72 | NE | NE |

| Recommended sleep duration<br>(≥7 hours) vs. Short (<7 hours) |  | Few insomnia symptoms<br>(yes vs. no)<sup>a</sup> |  | Restorative sleep<br>(yes vs. no)<sup>b</sup> |  |
| --- | --- | --- | --- | --- | --- |
| Model 1<sup>c</sup> | Model 2<sup>c</sup> | Model 1<sup>c</sup> | Model 2<sup>c</sup> | Model 1<sup>c</sup> | Model 2<sup>c</sup> |
| Prevalence Ratio (95% Confidence Interval) for Associations with Life satisfaction<sup>d</sup> (satisfied vs. dissatisfied) |  |  |  |  |  |
| (0.85 - 2.33) | (0.83 - 2.19) | (0.93 - 4.05) | (0.78 - 3.77) |  |  |

Abbreviations: NH (non-Hispanic), NE (not able to estimate)
<sup>a</sup>Few insomnia defined as not having trouble falling and staying asleep 'most days' or 'every day' in the past 30 days
<sup>b</sup>Restorative sleep defined as feeling rested upon waking 'most days' or 'every day' in the past 30 days
<sup>c</sup>Model 1 is adjusted for age (years) when not stratified by age, sex (men, women), student status (undergraduate students, graduate students), marital status (divorced/widowed/separated/married-spouse absent, single/never married, married-spouse present/living with partner), ratio of family income to poverty threshold (<1.00, 1.00-1.99, 2.00-2.99, ≥3.00), employment status (employed, not employed), and region of residence (Northeast, Midwest, South, West). Model 2 is adjusted for covariates in Model 1 and BMI (underweight (<18.5 kg/m)/recommended (18.5-24.9 kg/m), overweight (25-29.9 kg/m), obese (≥30 kg/m)), leisure-time physical activity (inactive, insufficiently active, sufficiently active), smoking status (never/quit > 12 months prior to interview, former, current), alcohol consumption (current, former, lifetime abstainer), and depression (yes, no). Models in the overall sample are additionally adjusted for race and ethnicity (Hispanic/Latine, NH-Black/African American, NH-Other, NH-White). NH-Other includes non-Hispanic, Asian, American Indian/Alaska native, Multiple and other races.
<sup>d</sup>Life satisfaction defined as 'satisfied' or 'very satisfied' vs. 'dissatisfied' or 'very dissatisfied' to the survey question 'In general, how satisfied are you with your life? Are you very satisfied, satisfied, dissatisfied, or very dissatisfied?'
All estimates are weighted for the complex survey design. Bolded values indicate statistical significance at a two-sided p-value < .05

**Table 3.** Prevalence ratios for associations between life satisfaction and sleep among college students, overall and stratified by race and ethnicity and sex, National Health Interview Survey, 2022, (N=1,426)

|  | Recommended sleep duration<br>(≥7 hours) vs. short (<7 hours) |  | Few insomnia symptoms<br>(yes vs. no)<sup>a</sup> |  | Restorative sleep<br>(yes vs. no)<sup>b</sup> |  |
| --- | --- | --- | --- | --- | --- | --- |
|  | Model 1<sup>c</sup> | Model 2<sup>c</sup> | Model 1<sup>c</sup> | Model 2<sup>c</sup> | Model 1<sup>c</sup> | Model 2<sup>c</sup> |
|  | Prevalence Ratio (95% Confidence Interval) for Associations with Life satisfaction<sup>d</sup> (satisfied vs. dissatisfied) |  |  |  |  |  |
| **Overall (n=1,426)** | **2.09**<br>**(1.28 - 3.42)** | **1.96**<br>**(1.19 - 3.22)** | **2.77**<br>**(1.51 - 5.08)** | **2.48**<br>**(1.36 - 4.52)** | **4.43**<br>**(1.77 - 11.1)** | **3.62**<br>**(1.49 - 8.83)** |
| Men (n=582) | **2.48**<br>**(1.08 - 5.68)** | **2.46**<br>**(1.09 - 5.57)** | **3.03**<br>**(1.29 - 7.12)** | **2.94**<br>**(1.27 - 6.78)** | **3.10**<br>**(1.14 - 8.41)** | **2.83**<br>**(1.07 - 7.47)** |
| Women (n=844) | 1.68<br>(0.98 - 2.88) | 1.43<br>(0.80 - 2.56) | **2.36**<br>**(1.16 - 4.78)** | 1.84<br>(0.88 - 3.83) | **15.8**<br>**(2.21 - 112.8)** | **10.1**<br>**(1.47 - 69.4)** |
| **Hispanic/Latine (n=299)** | **3.21**<br>**(1.12 - 9.21)** | 3.07<br>(0.96 - 9.86) | **7.89**<br>**(1.86 - 33.5)** | **7.29**<br>**(1.61 - 33.1)** | **5.58**<br>**(1.36 - 22.9)** | **4.86**<br>**(1.14 - 20.7)** |
| Men (n=111) | 7.89<br>(0.96 - 64.7) | 8.07<br>(0.96 - 67.7) | NE | NE | **7.19**<br>**(1.02 - 50.9)** | 7.54<br>(0.94 - 60.6) |
| Women (n=188) | 1.72<br>(0.84 - 3.53) | 1.10<br>(0.41 - 2.94) | 2.72<br>(0.81 - 9.08) | 1.90<br>(0.65 - 5.52) | 4.79<br>(0.65 - 35.4) | 1.68<br>(0.36 - 7.88) |
| **NH-Black/African American (n=200)** | 1.67<br>(0.43 - 6.45) | 1.93<br>(0.44 - 8.34) | 1.75<br>(0.63 - 4.84) | 1.76<br>(0.66 - 4.68) | 1.25<br>(0.37 - 4.22) | 1.17<br>(0.33 - 4.17) |
| Men (n=67) | 0.95<br>(0.36 - 2.51) | 0.93<br>(0.31 - 2.78) | 1.44<br>(0.66 - 3.14) | 1.45<br>(0.68 - 3.11) | 0.83<br>(0.24 - 2.90) | 1.10<br>(0.26 - 4.66) |
| Women (n=133) | NE | NE | NE | NE | NE | NE |
| **NH-Other (n=200)** | NE | NE | 4.34<br>(0.61 - 30.7) | 3.61<br>(0.56 - 23.4) | NE | NE |
| Men (n=89) | NE | NE | NE | NE | NE | NE |
| Women (n=111) | NE | NE | 3.37<br>(0.44 - 26.1) | 2.69<br>(0.42 - 17.3) | NE | NE |
| **NH-White (n=727)** | **1.72**<br>**(1.03 - 2.87)** | 1.63<br>(0.98 - 2.72) | **2.32**<br>**(1.08 - 4.96)** | 2.01<br>(0.92 - 4.40) | **6.74**<br>**(1.16 - 39.3)** | 5.25<br>(0.97 - 28.5) |
| Men (n=315) | 2.31<br>(0.85 - 6.25) | 2.32<br>(0.94 - 5.72) | 2.85<br>(0.97 - 8.34) | 2.67<br>(0.93 - 7.63) | 4.26<br>(0.73 - 24.7) | 3.34<br>(0.67 - 16.7) |
| Women (n=412) | 1.24 | 1.09 | 1.71 | 1.41 | NE | NE |

| Recommended sleep duration<br>(≥7 hours) vs. short (<7 hours) |  | Few insomnia symptoms<br>(yes vs. no)<sup>a</sup> |  | Restorative sleep<br>(yes vs. no)<sup>b</sup> |  |
| --- | --- | --- | --- | --- | --- |
| Model 1<sup>c</sup> | Model 2<sup>c</sup> | Model 1<sup>c</sup> | Model 2<sup>c</sup> | Model 1<sup>c</sup> | Model 2<sup>c</sup> |
| Prevalence Ratio (95% Confidence Interval) for Associations with Life satisfaction<sup>d</sup> (satisfied vs. dissatisfied) |  |  |  |  |  |
| (0.69 - 2.24) | (0.55 - 2.16) | (0.71 - 4.14) | (0.52 - 3.87) |  |  |

Abbreviations: NH (non-Hispanic), NE (not able to estimate)
<sup>a</sup>Few insomnia defined as not having trouble falling and staying asleep 'most days' or 'every day' in the past 30 days
<sup>b</sup>Restorative sleep defined as feeling rested upon waking 'most days' or 'every day' in the past 30 days
<sup>c</sup>Model 1 is adjusted for age (years), sex (men, women) when not stratified by sex, student status (undergraduate students, graduate students), marital status (divorced/widowed/separated/married-spouse absent, single/never married, married-spouse present/living with partner), ratio of family income to poverty threshold (<1.00, 1.00-1.99, 2.00-2.99, ≥3.00), employment status (employed, not employed), and region of residence (Northeast, Midwest, South, West). Model 2 is adjusted for covariates in Model 1 and BMI (underweight (<18.5 kg/m)/recommended (18.5-24.9 kg/m), overweight (25-29.9 kg/m), obese (≥30 kg/m)), leisure-time physical activity (inactive, insufficiently active, sufficiently active), smoking status (never/quit > 12 months prior to interview, former, current), alcohol consumption (current, former, lifetime abstainer), and depression (yes, no). Models in the overall sample are additionally adjusted for race and ethnicity (Hispanic/Latine, NH-Black/African American, NH-Other, NH-White). NH-Other includes non-Hispanic, Asian, American Indian/Alaska native, Multiple and other races.
<sup>d</sup>Life satisfaction defined as 'satisfied' or 'very satisfied' vs. 'dissatisfied' or 'very dissatisfied' to the survey question 'In general, how satisfied are you with your life? Are you very satisfied, satisfied, dissatisfied, or very dissatisfied?'
All estimates are weighted for the complex survey design. Bolded values indicate statistical significance at a two-sided p-value < .05

### Associations between life satisfaction and sleep by age, overall and within racial and ethnic groups

Age did not modify the overall associations with recommended sleep duration, few IS, nor restorative sleep either in the overall sample or within racial and ethnic groups (*p* interaction >0.05) (Table 2).

### Associations between life satisfaction and sleep by sex, overall and within racial and ethnic groups

Sex did not modify the overall associations with recommended sleep duration, few IS, nor restorative sleep (Table 3). In exploratory analyses, sex did not modify the associations within race and ethnicity groups.

### Associations between life satisfaction and sleep by race and ethnicity

Race and ethnicity did not modify associations with recommended sleep duration, few IS, and restorative sleep in the overall population (all *p* for cross-product terms >0.05) (Tables 2-3).

### Sensitivity analysis: Associations between life satisfaction and sleep by student status, overall and within racial and ethnic groups

Detailed age-standardized distributions of sociodemographic, health behavior, and clinical characteristics, overall and stratified by student status, are presented in Supplemental Table 6. Overall and within racial and ethnic groups, the unadjusted prevalence of life satisfaction generally did not differ by student status with one exception: among NH-Black/African American participants, life satisfaction differed by student status (p=0.031), with a higher prevalence among undergraduate students (99.5%) than among graduate students (98.0%) (Supplemental Table 7). This finding should be interpreted with caution because of the small sample size. Student status did not modify the associations with recommended sleep duration, few IS, or restorative sleep overall or within racial and ethnic groups (Table 4).

**Table 4.** Prevalence ratios for associations between life satisfaction and sleep among college students, overall and stratified by race and ethnicity and the highest educational attainment, National Health Interview Survey, 2022, (N=1,426)

|  | Recommended sleep duration<br>(≥7 hours) vs. short (<7 hours) |  | Few insomnia symptoms<br>(yes vs. no)<sup>a</sup> |  | Restorative sleep<br>(yes vs. no)<sup>b</sup> |  |
| --- | --- | --- | --- | --- | --- | --- |
|  | Model 1<sup>c</sup> | Model 2<sup>c</sup> | Model 1<sup>c</sup> | Model 2<sup>c</sup> | Model 1<sup>c</sup> | Model 2<sup>c</sup> |
|  | Prevalence Ratio (95% Confidence Interval) for Associations with Life satisfaction<sup>d</sup> (satisfied vs. dissatisfied) |  |  |  |  |  |
| **Overall (n=1,426)** | **2.09**<br>**(1.28 - 3.42)** | **1.96**<br>**(1.19 - 3.22)** | **2.77**<br>**(1.51 - 5.08)** | **2.48**<br>**(1.36 - 4.52)** | **4.43**<br>**(1.77 - 11.1)** | **3.62**<br>**(1.49 - 8.83)** |
| Undergraduate students (n=898) | **2.48**<br>**(1.26 - 4.84)** | **2.34**<br>**(1.20 - 4.59)** | **3.18**<br>**(1.44 - 7.00)** | **2.89**<br>**(1.33 - 6.27)** | **4.41**<br>**(1.45 - 13.4)** | **3.78**<br>**(1.27 - 11.2)** |
| Graduate students (n=528) | 1.39<br>(0.90 - 2.16) | 1.26<br>(0.79 - 2.00) | 1.74<br>(0.77 - 3.94) | 1.38<br>(0.60 - 3.20) | **4.69**<br>**(1.29 - 17.1)** | **3.33**<br>**(1.11 - 10.0)** |
| **Hispanic/Latine (n=299)** | **3.21**<br>**(1.12 - 9.21)** | 3.07<br>(0.96 - 9.86) | **7.89**<br>**(1.86 - 33.5)** | **7.29**<br>**(1.61 - 33.1)** | **5.58**<br>**(1.36 - 22.9)** | **4.86**<br>**(1.14 - 20.7)** |
| Undergraduate students (n=239) | 6.55<br>(0.94 - 45.6) | 7.03<br>(0.95 - 52.2) | **14.5**<br>**(1.86 - 112.9)** | **14.1**<br>**(1.79 - 111.1)** | **9.69**<br>**(1.23 - 76.6)** | **9.25**<br>**(1.14 - 74.7)** |
| Graduate students (n=60) | NE | NE | 1.62<br>(0.39 - 6.84) | NE | 1.74<br>(0.58 - 5.25) | NE |
| **NH-Black/African American (n=200)** | 1.67<br>(0.43 - 6.45) | 1.93<br>(0.44 - 8.34) | 1.75<br>(0.63 - 4.84) | 1.76<br>(0.66 - 4.68) | 1.25<br>(0.37 - 4.22) | 1.17<br>(0.33 - 4.17) |
| Undergraduate students (n=129) | 1.64<br>(0.45 - 5.98) | 1.69<br>(0.42 - 6.76) | 1.91<br>(0.58 - 6.26) | 1.87<br>(0.59 - 5.90) | 1.49<br>(0.33 - 6.62) | 1.40<br>(0.29 - 6.82) |
| Graduate students (n=71) | NE | NE | NE | NE | NE | NE |
| **NH-Other (n=200)** | NE | NE | 4.34<br>(0.61 - 30.7) | 3.61<br>(0.56 - 23.4) | NE | NE |
| Undergraduate students (n=115) | NE | NE | 2.31<br>(0.43 - 12.4) | 2.43<br>(0.50 - 11.9) | NE | NE |
| Graduate students (n=85) | NE | NE | NE | NE | NE | NE |
| **NH-White (n=727)** | **1.72**<br>**(1.03 - 2.87)** | 1.63<br>(0.98 - 2.72) | **2.32**<br>**(1.08 - 4.96)** | 2.01<br>(0.92 - 4.40) | **6.74**<br>**(1.16 - 39.3)** | 5.25<br>(0.97 - 28.5) |
| Undergraduate students (n=415) | 2.03<br>(0.99 - 4.15) | **1.97**<br>**(1.01 - 3.83)** | **2.79**<br>**(1.02 - 7.63)** | 2.41<br>(0.89 - 6.54) | 5.35<br>(0.98 - 29.1) | 4.21<br>(0.85 - 20.8) |
| Graduate students (n=312) | 1.35<br>(0.80 - 2.27) | 1.27<br>(0.68 - 2.38) | 1.56<br>(0.63 - 3.87) | 1.14<br>(0.42 - 3.09) | NE | NE |

| Recommended sleep duration<br>(≥7 hours) vs. short (<7 hours) |  | Few insomnia symptoms<br>(yes vs. no)<sup>a</sup> |  | Restorative sleep<br>(yes vs. no)<sup>b</sup> |  |
| --- | --- | --- | --- | --- | --- |
| Model 1<sup>c</sup> | Model 2<sup>c</sup> | Model 1<sup>c</sup> | Model 2<sup>c</sup> | Model 1<sup>c</sup> | Model 2<sup>c</sup> |
| Prevalence Ratio (95% Confidence Interval) for Associations with Life satisfaction<sup>d</sup> (satisfied vs. dissatisfied) |  |  |  |  |  |

Abbreviations: NH (non-Hispanic), NE (not able to estimate)
<sup>a</sup> Few insomnia defined as not having trouble falling and staying asleep 'most days' or 'every day' in the past 30 days
<sup>b</sup> Restorative sleep defined as feeling rested upon waking 'most days' or 'every day' in the past 30 days

## DISUSSION

In this nationally-representative sample of U.S. college students, the prevalence of life satisfaction was 97.1% and did not vary significantly by age, sex, race, nor ethnicity. Students reporting life satisfaction had a significantly higher prevalence of recommended sleep duration, few IS, and restorative sleep, compared to students reporting life dissatisfaction. Neither age, sex, nor race along with ethnicity modified overall associations. Likewise, in exploratory analyses within racial and ethnic groups, results for modification of the life satisfaction-sleep relationship by sex and age yielded no evidence, which should be interpreted with caution due to small sample sizes within some subgroups. Furthermore, the associations between life satisfaction and sleep were generally consistent across undergraduate and graduate students.

The prevalence of life satisfaction among U.S. college students in our study was 97%, which was broadly consistent with the generally favorable levels of life satisfaction reported in previous national estimates among U.S. adults.^53^ However, to our knowledge, no prior studies have reported the prevalence of life satisfaction among U.S. college students using a comparable measure, limiting direct comparisons with previous college-based studies. One study of 12,646 Chinese college students assessed life satisfaction using the Satisfaction With Life Scale (SWLS) and reported a mean score of 23 out of 30.^48^ Another study of 254 undergraduate students in Israel assessed overall life satisfaction or quality of life using a 0-100 numerical rating scale and reported a mean score of 75.^49^ Differences in study populations and measurement approaches, however, limit direct comparison of life satisfaction levels across studies.

Our findings that life satisfaction was associated with better sleep quality are consistent with prior literature. Specifically, studies of college students in Israel and China similarly showed positive correlations between life satisfaction and sleep quality.^48, 49^ We extend previous studies by using a nationally sample of U.S. college students while determining if findings differ across sociodemographic groups, thereby addressing an important gap in the U.S. literature.

The observed associations may be explained by individual and environmental-level factors. Life satisfaction is associated with greater psychological resilience and better mental health, which may contribute to fewer sleep disturbances by promoting more adaptive coping with stress and reducing stress-related psychological arousal that can interfere with sleep.^63^ Consistent with this potential pathway, prior research among college students has shown that mental health difficulties, including depression and anxiety, are associated with lower life satisfaction and may also contribute to sleep disturbances.^64^ Although factors that may contribute to life satisfaction were not directly measured in the present study, several potential social and environmental pathways may help explain the observed association between life satisfaction and sleep. For example, greater campus involvement and a stronger sense of belonging are associated with better mental health and more regular sleep patterns.^9, 65^ Conversely, a lack of belonging could disrupt both life satisfaction and sleep. Overall, greater life satisfaction may reflect psychological and social resources that help students cope with college-related stressors, including challenges related to social integration and belonging, which may, in turn, be associated with fewer sleep disturbances.^66^

Despite the lack of evidence in this study, differences by sociodemographic characteristics remain plausible. For instance, differences by sex may stem from underlying biological, psychological, and social differences in stress processing, emotional processes, and sleep vulnerability.^67^ Women’s sleep may be more susceptible to hormonal fluctuations (e.g., menstrual cycle), which can disrupt circadian rhythms and sleep architecture,^68^ potentially attenuating the association between life satisfaction and sleep.^69^ Women, for instance, may face unique social stressors in academic settings—based on expectations or pressures—that may not be as readily alleviated by general life satisfaction.^70^ In turn, men may have a more efficient stress regulation system, which allows positive emotions—linked to life satisfaction—to more effectively buffer against sleep disruptions.^71^ However, male students may be more likely to underreport poor sleep health issues ^8^ and may also be less likely to express vulnerability, acknowledge psychological distress, or seek help.^72^ Similarly, reporting and coping patterns may influence differences in associations by age, race, and ethnicity among college students.^73, 74^ Further research with larger samples is warranted to evaluate potential modification of the life satisfaction-sleep relationship by age, sex, race and ethnicity.

In sensitivity analyses, we found no evidence that the associations between life satisfaction and sleep differed by student status (undergraduate vs graduate; however, the absence of significant differences may partly reflect limited statistical power due to reduced sample sizes after stratification. Nevertheless, differences by student status remain plausible given the distinct academic, social, and occupational demands experienced by undergraduate and graduate students.^75^ Undergraduate students may face academic pressures, evolving social roles, and less-established daily routines that can affect sleep,^76^ whereas graduate students may experience different stressors, including greater career-related demands and competing professional or personal responsibilities.^76, 77^ Further research across groups of undergraduate and graduate students is warranted.

The cross-sectional design limits knowledge of the direction of associations and limits causal inference between life satisfaction and sleep. The survey period may also have been influenced by the residual effects of the COVID-19 pandemic, which could have impacted college students’ life satisfaction and overall health. Although the sample was nationally representative, there were small sample sizes of many racial and ethnic groups, especially upon additional stratification by age and sex, limiting power and precision. Our measure of life satisfaction in this study was based on a self-reported question, which may be subject to reporting bias and potential misclassification, as responses may be influenced by current mood or individual interpretation of the question as individuals may have unique criteria for values and goals that affect satisfaction.^78^ Similar limitations apply to the assessment of sleep characteristics, which were also self-reported and may, therefore, be subject to recall bias and measurement error.^79, 80^ However, this approach is practical for large-scale national surveys and provides a direct measure of perceived life satisfaction. Future studies could use validated instruments such as the Satisfaction With Life Scale (SWLS),^81^ which has demonstrated discriminant validity from measures of emotional well-being. The SWLS is widely used and allows for more consistent comparisons across demographical groups, as well as exploration of specific components contributing to life satisfaction.^82^ Lastly, we were unable to capture temporal changes in life satisfaction and sleep outcomes. Given that college students experience key college-related transitions (e.g. transitioning from early college, when academic demands are the primary focus, to later stages approaching graduation, when concerns about future career and life direction may increase), future longitudinal studies incorporating detailed information on life events with repeated measures are needed to better understand the potential bidirectional associations between life satisfaction and sleep among this group.

Despite these limitations, our data were collected from the 2022 NHIS, which provided a relatively large and representative sample of college students in the U.S. We investigated the relationship between life satisfaction and sleep disturbances overall and highlighted differences across age, sex, and race and ethnicity - sociodemographic factors that have largely not been assessed as modifiers in prior research.^48, 49^ Our analysis also accounted for multiple potential confounders, thereby strengthening the validity of our findings. In addition, we captured multiple measures of sleep health, including sleep duration, IS, and restorative sleep.^58^

Our study highlights the potential importance of holistic well-being initiatives on college campuses. Interventions aimed at improving students’ overall life satisfaction—through enhanced support for career planning, social connectedness, financial stability, physical health, and community engagement— and sleep may serve as effective strategies to promote overall well-being. Future research should investigate the life satisfaction-sleep relationship using longitudinal research designs with larger samples and objective sleep measures, which allows for investigation of causal relationships and potential changes in relationships over time. Qualitative interviews, as a complementary approach, may also help better identify sources of life stress that influence life satisfaction, which aid the development of targeted strategies to promote sleep health.

Life satisfaction was prevalent and associated with favorable sleep health, including recommended sleep duration, few IS, and restorative sleep among U.S. college students. Although associations were similar across age, sex, racial, and ethnic groups, more research is warranted. Future studies should also further investigate factors that support life satisfaction as an opportunity to improve or maintain sleep health along with the role of favorable sleep health in the promotion of life satisfaction.

## Conflict of interest disclosure

The authors have no conflicts of interest to report.

## Availability of data and materials

The datasets analyzed during the current study are publicly available from the National Center for Health Statistics (NCHS), Centers for Disease Control and Prevention, National Health Interview Survey (NHIS), at: https://www.cdc.gov/nchs/nhis/index.htm

## Acknowledgements

The authors wish to thank the National Health Interview Survey (NHIS) participants. The datasets generated during and/or analyzed during the current study are publicly available.

## AUTHOR CONTRIBUTIONS

*Authors:* Bethany T. Ogbenna, Wensu Zhou, LaDarius T. Williams, Symielle A. Gaston, Philip M. Zendels, Christopher Payne, W. B. Jackson II, Chandra L. Jackson

*Study concept:* CL. Jackson.

*Study design:* CL Jackson, BT. Ogbenna.

*Acquisition of data:* C. Payne.

*Statistical Analysis:* C. Payne.

*Interpretation of data:* BT. Ogbenna, W. Zhou, SA. Gaston, C. Payne, WB. Jackson II, CL. Jackson.

*Drafting of the manuscript:* BT. Ogbenna, W. Zhou, LT. Williams

*Critical revision of the manuscript for important intellectual content:* BT. Ogbenna, W. Zhou, LT. Williams, SA. Gaston, PM Zendels, C Payne, WB Jackson II, CL. Jackson.

*Administrative, technical, and material support:* CL. Jackson, SA. Gaston,

*Obtaining funding:* CL. Jackson.

*Study supervision:* CL. Jackson, WB. Jackson II.

*Final Approval:* BT. Ogbenna, W. Zhou, LT. Williams, SA. Gaston, PM. Zendels, C. Payne, WB. Jackson II, CL. Jackson.

## Notes

Sources of funding: This research was supported, in part, by the Intramural Research Program of the NIH, National Institute of Environmental Health Sciences (Z1A ES103325 [CLJ]). The contributions of the NIH authors are considered Works of the United States Government. The findings and conclusions presented in this paper are those of the authors and do not necessarily reflect the views of the NIH or the U.S. Department of Health and Human Services.

### Competing Interest Statement

The authors have declared no competing interest.

