## Supplemental Material for "Life Satisfaction and Sleep Health among College Students in the United States"

### Supplemental Figure 1. Flow chart of study population selection

Final analytic sample of adult college students

**N=1,426**

- Missing data
  - All sleep measures (n=5)
  - Life satisfaction (n=3)
  - Sex (n=1)
  - Racial and ethnic groups (n=0)
  - Age (n=0)
  - Potential confounders (n=23)
- **Total excluded (n=32)**

Sample of adults not currently attending school and/or have not completed high school/GED equivalent or more (n = 26,193)

NHIS Sample of college students, 2022

**(N=1,458)**

NHIS Sample of adults,

2022

**(N=27,651)**

### Supplemental Figure 2. Directed acyclic graph for the association between life satisfaction and sleep disturbances

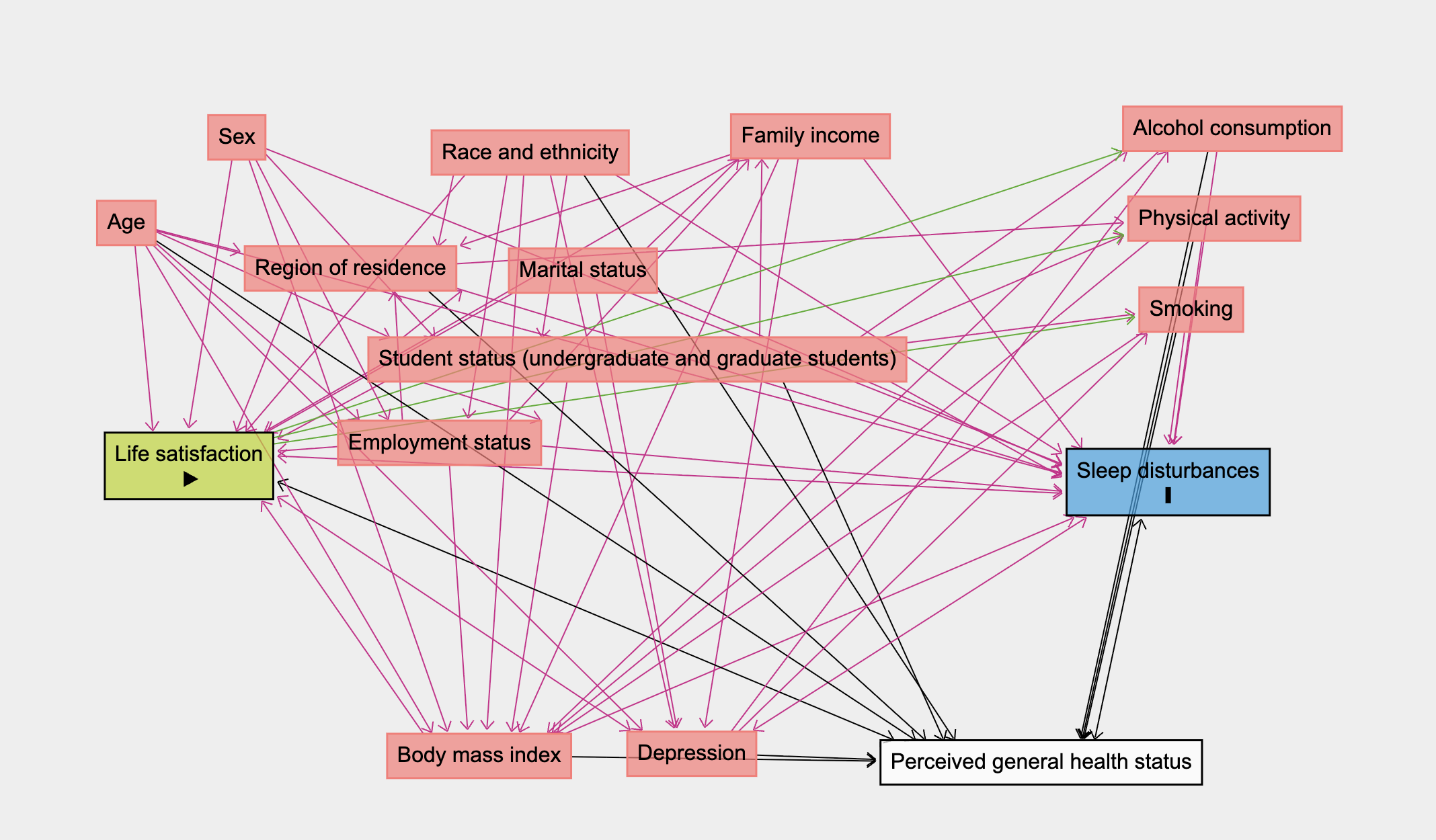

### Supplemental Table 1. Comparison of included and excluded study participants, National Health Interview Survey, 2022 (N=1,458)

|  | Included  n=1,426 (97.8%) | Excluded  n=32 (2.2%) | Chi-square or  t-test p-value |
| --- | --- | --- | --- |
| **Sociodemographic Characteristics** |  |  |  |
| Age (years), mean (SE) | 27.7 (0.3) | 24.0 (1.7) | 0.036 |
| Age group |  |  | 0.019 |
| <23 years | 45.8 | 67.4 |  |
| ≥23 years | 54.2 | 32.6 |  |
| Sex* |  |  | 0.857 |
| Men | 38.6 | 29.2 |  |
| Women | 61.4 | 70.8 |  |
| Race/ethnicity |  |  | 0.062 |
| Hispanic/Latine | 14.8 | 11.6 |  |
| NH-Black/African American | 13.2 | 21.1 |  |
| NH-Other | 10.6 | 43.7 |  |
| NH-White | 61.4 | 23.6 |  |
| Student status |  |  | <0.001 |
| Undergraduate students | 58.5 | 91.2 |  |
| Graduate students | 41.5 | 8.8 |  |
| Employment status* |  |  | 0.197 |
| Employed | 59.5 | 36.5 |  |
| Not employed | 40.5 | 63.5 |  |
| Ratio of family income to poverty threshold |  |  | 0.115 |
| <1.00 | 7.8 | 11.0 |  |
| 1.00-1.99 | 15.1 | 48.5 |  |
| 2.00-2.00 | 16.0 | 10.7 |  |
| ≥3.00 | 61.1 | 29.8 |  |
| Life satisfaction (yes)* ^a^ | 98.1 | 100.0 | <0.001 |
| Marital status* |  |  | 0.173 |
| Divorced/widowed/separated/married-spouse absent | 18.7 | 42.4 |  |
| Single/never married | 36.9 | 45.6 |  |
| Married-spouse present/living with partner | 44.4 | 12.0 |  |
| Region of residence |  |  | 0.008 |
| Northeast | 16.2 | 54.3 |  |
| Midwest | 22.0 | 12.8 |  |
| South | 34.7 | 25.2 |  |
| West | 27.1 | 7.7 |  |
| **Health Behaviors** |  |  |  |
| Smoking status* |  |  | 0.724 |
| Never/quit >12 months prior | 95.0 | 100.0 |  |
| Former/quit≤12 months ago | 0.8 | 0.0 |  |
| Current | 4.2 | 0.0 |  |
| Alcohol consumption* |  |  | 0.005 |
| Current (≥1 drink past year) | 67.0 | 41.5 |  |
| Former (no drinks past year) | 17.2 | 3.7 |  |
| Lifetime abstinence (<12 drinks in life) | 15.8 | 54.8 |  |
| Leisure-time physical activity* |  |  | 0.277 |
| Inactive | 18.0 | 45.7 |  |
| Insufficiently active | 25.2 | 13.5 |  |
| Sufficiently active | 56.8 | 40.8 |  |
| Usual sleep duration* |  |  | 0.153 |
| Short (<7 hours) | 29.3 | 9.9 |  |
| Recommended (≥7 hours) | 70.7 | 90.1 |  |
| Few insomnia (yes)* ^b^ | 76.6 | 48.0 | <0.001 |
| Restorative sleep (yes)* ^c^ | 61.0 | 35.5 | 0.004 |
| **Clinical Characteristics** |  |  |  |
| Ever had depression (yes)* ^d^ | 23.2 | 9.2 | 0.004 |
| Body mass index category* |  |  | 0.289 |
| Underweight (<18.5 kg/m^2^) | 1.5 | 5.8 |  |
| Recommended (18.5-<25 kg/m^2^) | 36.8 | 39.6 |  |
| Overweight (25-<30 kg/m^2^) | 32.7 | 54.6 |  |
| Obesity (≥30 kg/m^2^) | 28.9 | 0.0 |  |
| General health status |  |  | 0.009 |
| Excellent/Very good/Good | 93.1 | 56.6 |  |
| Fair/poor | 6.9 | 43.4 |  |
| Abbreviations: SE (standard error), NH (non-Hispanic) | | | |
| Note: All estimates are age-standardized to the US 2020 population, except for age and weighted for the survey's complex sampling design. Data are presented as column percentages or means and standard errors. Percentages may not sum to 100 due to rounding or missing data. College students are defined as having completed High School or more (EDUC) and currently attending school (SCHOOLNOW) | | | |
| ^a^ Life satisfaction defined as a response of 'very dissatisfied' or 'dissatisfied' vs. 'satisfied' or 'very satisfied'. 'In general, how satisfied are you with your life? Are you very satisfied, satisfied, dissatisfied, or very dissatisfied?' | | | |
| ^b^ Few insomnia defined as not having trouble falling and staying asleep 'most days' or 'every day' in past 30 days | | | |
| ^c^ Restorative sleep defined as feeling rested upon waking 'most days' or 'every day' in past 30 days | | | |
| ^d^ Depression was defined by the 2019 Field Representative's Manual defines depression (major depressive disorder or clinical depression) as a common but serious mood disorder. It causes severe symptoms that affect how you feel, think, and handle daily activities, such as sleeping, eating, or working. | | | |
| * We excluded participants with missing data on confounders from the prevalence estimates and statistical tests. | | | |

### Supplemental Table 2. Prevalence of life satisfaction among college students by race and ethnicity and age, National Health Interview Survey, 2022, (N=1,426)

|  | Total | | | | Hispanic/Latine | | | | NH-Black/African American | | | | NH-Other^*^ | | | | NH-White | | | |
| --- | --- | --- | --- | --- | --- | --- | --- | --- | --- | --- | --- | --- | --- | --- | --- | --- | --- | --- | --- | --- |
|  | All n=1,426 (100%) | <23 years n=457 (32.0%) | ≥23 years n=969 (68.0%) | Chi-square  or t-test  p-value | All n=299 (100%) | <23 years n=116 (38.8%) | ≥23 years n=183 (61.2%) | Chi-square  or t-test  p-value | All n=200 (100%) | <23 years n=47 (23.5%) | ≥23 years n=153 (76.5%) | Chi-square  or t-test  p-value | All n=200 (100%) | <23 years n=80 (40.0%) | ≥23 years n=120 (60.0%) | Chi-square  or t-test  p-value | All n=727 (100%) | <23 years n=214 (29.4%) | ≥23 years n=513 (70.6%) | Chi-square  or t-test  p-value |
| Life satisfaction ^a^ |  |  |  | 0.465 |  |  |  | 0.551 |  |  |  | 0.061 |  |  |  | 0.332 |  |  |  | 0.900 |
| Dissatisfied | 1.9 | 2.4 | 2.1 |  | 2.7 | 3.5 | 2.5 |  | 1.6 | 5.0 | 1.2 |  | 0.6 | 0.0 | 1.3 |  | 2.1 | 2.0 | 2.4 |  |
| Satisfied | 98.1 | 97.6 | 97.9 |  | 97.3 | 96.5 | 97.5 |  | 98.4 | 95.0 | 98.8 |  | 99.4 | 100.0 | 98.7 |  | 97.9 | 98.0 | 97.6 |  |
| Abbreviations: NH (non-Hispanic) | | | | | | | | | | | | | | | | | | | | |
| Note: All estimates are age-standardized to the US 2020 population, except for age and weighted for the survey's complex sampling design. Data are presented as column percentages or means and standard errors. Percentages may not sum to 100 due to rounding or missing data. College students are defined as having completed High School or more (EDUC) and currently attending school (SCHOOLNOW) | | | | | | | | | | | | | | | | | | | | |
| ^a^ Life satisfaction defined as a response of 'very dissatisfied' or 'dissatisfied' vs. 'satisfied' or 'very satisfied'. 'In general, how satisfied are you with your life? Are you very satisfied, satisfied, dissatisfied, or very dissatisfied?' | | | | | | | | | | | | | | | | | | | | |
| ^*^ NH-Other includes non-Hispanic, Asian, American Indian/Alaska native, Multiple and other races. | | | | | | | | | | | | | | | | | | | | |

### Supplemental Table 3. Prevalence of life satisfaction among college students by race and ethnicity and sex, National Health Interview Survey, 2022, (N=1,426)

|  | Total | | | | Hispanic/Latine | | | | NH-Black/African American | | | | NH-Other^*^ | | | | NH-White | | | |
| --- | --- | --- | --- | --- | --- | --- | --- | --- | --- | --- | --- | --- | --- | --- | --- | --- | --- | --- | --- | --- |
|  | All n=1,426 (100%) | Men n=582 (40.8%) | Women n=844 (59.2%) | Chi-square  or t-test  p-value | All n=299 (100%) | Men n=111 (37.1%) | Women n=188 (62.9%) | Chi-square  or t-test  p-value | All n=200 (100%) | Men n=67 (33.5%) | Women n=133 (66.5%) | Chi-square  or t-test  p-value | All n=200 (100%) | Men n=89 (44.5%) | Women n=111 (55.5%) | Chi-square  or t-test  p-value | All n=727 (100%) | Men n=315 (43.3%) | Women n=412 (56.7%) | Chi-square  or t-test  p-value |
| Life satisfaction ^a^ |  |  |  | 0.136 |  |  |  | 0.298 |  |  |  | 0.058 |  |  |  | 0.924 |  |  |  | 0.521 |
| Dissatisfied | 1.9 | 2.4 | 1.5 |  | 2.7 | 3.0 | 2.0 |  | 1.6 | 3.0 | 0.7 |  | 0.6 | 0.5 | 0.7 |  | 2.1 | 2.5 | 1.8 |  |
| Satisfied | 98.1 | 97.6 | 98.5 |  | 97.3 | 97.0 | 98.0 |  | 98.4 | 97.0 | 99.3 |  | 99.4 | 99.5 | 99.3 |  | 97.9 | 97.5 | 98.2 |  |
| Abbreviations: NH (non-Hispanic) | | | | | | | | | | | | | | | | | | | | |
| Note: All estimates are age-standardized to the US 2020 population, except for age and weighted for the survey's complex sampling design. Data are presented as column percentages or means and standard errors. Percentages may not sum to 100 due to rounding or missing data. College students are defined as having completed High School or more (EDUC) and currently attending school (SCHOOLNOW) | | | | | | | | | | | | | | | | | | | | |
| ^a^ Life satisfaction defined as a response of 'very dissatisfied' or 'dissatisfied' vs. 'satisfied' or 'very satisfied'. 'In general, how satisfied are you with your life? Are you very satisfied, satisfied, dissatisfied, or very dissatisfied?' | | | | | | | | | | | | | | | | | | | | |
| ^*^ NH-Other includes non-Hispanic, Asian, American Indian/Alaska native, Multiple and other races. | | | | | | | | | | | | | | | | | | | | |

### Supplemental Table 4. Age-standardized sociodemographic, health behavior, and clinical characteristics among college students, overall and by age, National Health Interview Survey, 2022 (N=1,426)

|  | Total  n=1,426 (100%) | | | | <23 years  n=457 (32.0%) | | | | ≥23 years  n=969 (68.0%) | | | |
| --- | --- | --- | --- | --- | --- | --- | --- | --- | --- | --- | --- | --- |
|  |  | Life satisfaction ^a^ | | Chi-square or t-test  p-value |  | Life satisfaction ^a^ | | Chi-square or t-test  p-value |  | Life satisfaction ^a^ | | Chi-square or t-test  p-value |
|  | All n=1,426 (100%) | Yes n=1,385 (97.1%) | No n=41 (2.9%) |  | All n=457 (100%) | Yes n=446 (97.6%) | No n=11 (2.4%) |  | All n=969 (100%) | Yes n=939 (96.9%) | No n=30 (3.1%) |  |
| **Sociodemographic Characteristics** |  | | | |  | | | |  | | | |
| Age (years), mean (SE) | 27.7 (0.3) | 27.7 (0.3) | 26.9 (1.3) | 0.566 | 19.8 (0.1) | 19.8 (0.1) | 19.9 (0.4) | 0.809 | 34.3 (0.4) | 34.4 (0.4) | 31.6 (1.6) | 0.095 |
| Sex |  |  |  | 0.136 |  |  |  | 0.349 |  |  |  | 0.140 |
| Men | 38.6 | 38.4 | 50.2 |  | 49.4 | 49.0 | 64.7 |  | 36.7 | 36.4 | 48.4 |  |
| Women | 61.4 | 61.6 | 49.8 |  | 50.6 | 51.0 | 35.3 |  | 63.3 | 63.6 | 51.6 |  |
| Race and ethnicity |  |  |  | 0.390 |  |  |  | 0.401 |  |  |  | 0.788 |
| Hispanic/Latine | 14.8 | 14.7 | 20.8 |  | 23.2 | 22.9 | 34.5 |  | 14.2 | 14.2 | 17.3 |  |
| NH-Black/African American | 13.2 | 13.3 | 10.1 |  | 10.3 | 10.0 | 21.6 |  | 14.7 | 14.9 | 8.2 |  |
| NH-Other | 10.6 | 10.8 | 3.5 |  | 13.9 | 14.3 | 0.0 |  | 10.1 | 10.2 | 6.1 |  |
| NH-White | 61.4 | 61.2 | 65.6 |  | 52.6 | 52.8 | 43.9 |  | 60.9 | 60.7 | 68.3 |  |
| Student status |  |  |  | 0.273 |  |  |  | 0.467 |  |  |  | 0.264 |
| Undergraduate students | 58.5 | 58.5 | 65.5 |  | 94.9 | 94.7 | 100.0 |  | 49.9 | 49.8 | 57.9 |  |
| Graduate students | 41.5 | 41.5 | 34.5 |  | 5.1 | 5.3 | 0.0 |  | 50.1 | 50.2 | 42.1 |  |
| Employment status |  |  |  | 0.176 |  |  |  | 0.487 |  |  |  | 0.232 |
| Employed | 59.5 | 60.0 | 44.7 |  | 60.8 | 61.1 | 49.4 |  | 63.1 | 63.7 | 47.5 |  |
| Not employed | 40.5 | 40.0 | 55.3 |  | 39.2 | 38.9 | 50.6 |  | 36.9 | 36.3 | 52.5 |  |
| Ratio of family income to poverty threshold |  |  |  | 0.001 |  |  |  | 0.902 |  |  |  | <0.001 |
| <1.00 | 7.8 | 7.6 | 22.6 |  | 14.5 | 14.4 | 19.9 |  | 7.0 | 6.7 | 23.8 |  |
| 1.00-1.99 | 15.1 | 14.9 | 35.7 |  | 17.5 | 17.6 | 16.0 |  | 15.3 | 15.0 | 36.2 |  |
| 2.00-2.99 | 16.0 | 15.9 | 15.5 |  | 16.2 | 16.1 | 21.8 |  | 16.3 | 16.1 | 17.6 |  |
| ≥3.00 | 61.1 | 61.6 | 26.2 |  | 51.7 | 51.9 | 42.3 |  | 61.4 | 62.1 | 22.4 |  |
| Marital status |  |  |  | 0.002 |  |  |  | 0.040 |  |  |  | <0.001 |
| Divorced/Widowed/Separated/Married-spouse absent | 18.7 | 18.5 | 21.6 |  | 0.4 | 0.4 | 0.0 |  | 20.2 | 20.1 | 23.5 |  |
| Single/Never married | 36.9 | 36.5 | 66.0 |  | 95.5 | 96.0 | 77.3 |  | 25.5 | 24.8 | 64.8 |  |
| Married-spouse present/Living with partner | 44.4 | 44.9 | 12.4 |  | 4.1 | 3.6 | 22.7 |  | 54.3 | 55.2 | 11.7 |  |
| Region of residence |  |  |  | 0.715 |  |  |  | 0.739 |  |  |  | 0.833 |
| Northeast | 16.2 | 16.1 | 21.2 |  | 20.7 | 20.4 | 35.1 |  | 15.2 | 15.2 | 15.9 |  |
| Midwest | 22.0 | 22.0 | 24.2 |  | 21.0 | 21.0 | 17.3 |  | 21.4 | 21.4 | 23.7 |  |
| South | 34.7 | 35.0 | 24.0 |  | 31.5 | 31.7 | 22.7 |  | 36.6 | 36.8 | 27.0 |  |
| West | 27.1 | 26.9 | 30.6 |  | 26.8 | 26.8 | 24.9 |  | 26.8 | 26.6 | 33.4 |  |
| **Health Behaviors** |  | | | |  | | | |  | | | |
| Smoking status |  |  |  | 0.184 |  |  |  | 0.939 |  |  |  | 0.128 |
| Never/quit>12 months prior | 95.0 | 95.2 | 85.9 |  | 99.0 | 99.0 | 100.0 |  | 93.3 | 93.5 | 82.3 |  |
| Former/quit≤12 months ago | 0.8 | 0.8 | 2.3 |  | 0.2 | 0.2 | 0.0 |  | 1.2 | 1.2 | 4.1 |  |
| Current | 4.2 | 4.1 | 11.8 |  | 0.8 | 0.8 | 0.0 |  | 5.5 | 5.4 | 13.6 |  |
| Alcohol consumption |  |  |  | 0.445 |  |  |  | 0.391 |  |  |  | 0.633 |
| Current (≥1 drink past year) | 67.0 | 66.8 | 76.3 |  | 58.5 | 58.0 | 77.9 |  | 72.3 | 72.3 | 78.5 |  |
| Former (no drinks past year) | 17.2 | 17.2 | 10.3 |  | 5.2 | 5.3 | 0.0 |  | 18.2 | 18.2 | 13.0 |  |
| Lifetime abstinence (<12 drinks in life) | 15.8 | 15.9 | 13.4 |  | 36.3 | 36.6 | 22.1 |  | 9.5 | 9.5 | 8.5 |  |
| Leisure-time physical activity |  |  |  | 0.040 |  |  |  | 0.387 |  |  |  | 0.266 |
| Inactive | 18.0 | 17.6 | 41.0 |  | 13.3 | 13.0 | 28.0 |  | 18.3 | 18.0 | 37.6 |  |
| Insufficiently active | 25.2 | 25.2 | 24.2 |  | 19.6 | 19.7 | 15.6 |  | 26.3 | 26.4 | 26.2 |  |
| Sufficiently active | 56.8 | 57.2 | 34.8 |  | 67.1 | 67.4 | 56.4 |  | 55.3 | 55.6 | 36.2 |  |
| Usual sleep duration |  |  |  | 0.009 |  |  |  | 0.001 |  |  |  | 0.051 |
| Short (<7 hours) | 29.3 | 28.9 | 47.1 |  | 23.6 | 22.4 | 73.6 |  | 32.2 | 31.9 | 44.9 |  |
| Recommended (≥7 hours) | 70.7 | 71.1 | 52.9 |  | 76.4 | 77.6 | 26.4 |  | 67.8 | 68.1 | 55.1 |  |
| Few insomnia (yes) ^b^ | 76.6 | 77.3 | 37.4 | <0.001 | 80.1 | 81.3 | 29.5 | 0.001 | 74.7 | 75.6 | 36.3 | <0.001 |
| Restorative sleep (yes) ^c^ | 61.0 | 61.9 | 8.0 | <0.001 | 59.3 | 60.4 | 14.2 | <0.001 | 59.1 | 60.0 | 7.7 | <0.001 |
| **Clinical Characteristics** |  | | | |  | | | |  | | | |
| Ever had depression (yes) ^d^ | 23.2 | 22.4 | 68.5 | <0.001 | 17.9 | 17.4 | 38.6 | 0.201 | 25.1 | 24.2 | 71.7 | <0.001 |
| Body mass index category |  |  |  | 0.015 |  |  |  | 0.397 |  |  |  | 0.158 |
| Underweight (<18.5 kg/m^2^) | 1.5 | 1.4 | 9.6 |  | 4.9 | 4.6 | 16.7 |  | 0.5 | 0.5 | 5.4 |  |
| Recommended (18.5-<25 kg/m^2^) | 36.8 | 37.0 | 24.7 |  | 54.0 | 54.1 | 50.3 |  | 32.2 | 32.4 | 21.2 |  |
| Overweight (25-<30 kg/m^2^) | 32.7 | 32.6 | 39.5 |  | 23.2 | 23.3 | 20.4 |  | 34.5 | 34.3 | 42.9 |  |
| Obese (≥30 kg/m^2^) | 28.9 | 29.0 | 26.2 |  | 17.8 | 18.0 | 12.6 |  | 32.7 | 32.8 | 30.5 |  |
| General health status |  |  |  | <0.001 |  |  |  | 0.077 |  |  |  | <0.001 |
| Excellent/Very good/Good | 93.1 | 93.5 | 71.3 |  | 96.3 | 96.6 | 83.3 |  | 92.6 | 93.2 | 66.4 |  |
| Fair/poor | 6.9 | 6.5 | 28.7 |  | 3.7 | 3.4 | 16.7 |  | 7.4 | 6.8 | 33.6 |  |
| Abbreviations: SE (standard error), NH (non-Hispanic) | | | | | | | | | | | | |
| Note: All estimates are age-standardized to the US 2020 population, except for age and weighted for the survey's complex sampling design. Data are presented as column percentages or means and standard errors. Percentages may not sum to 100 due to rounding or missing data. College students are defined as having completed High School or more (EDUC) and currently attending school (SCHOOLNOW) | | | | | | | | | | | | |
| ^a^ Life satisfaction defined as a response of 'very dissatisfied' or 'dissatisfied' vs. 'satisfied' or 'very satisfied'. 'In general, how satisfied are you with your life? Are you very satisfied, satisfied, dissatisfied, or very dissatisfied?' | | | | | | | | | | | | |
| ^b^ Few insomnia defined as not having trouble falling and staying asleep 'most days' or 'every day' in past 30 days | | | | | | | | | | | | |
| ^c^ Restorative sleep defined as feeling rested upon waking 'most days' or 'every day' in past 30 days | | | | | | | | | | | | |
| ^d^ Depression was defined by the 2019 Field Representative's Manual defines depression (major depressive disorder or clinical depression) as a common but serious mood disorder. It causes severe symptoms that affect how you feel, think, and handle daily activities, such as sleeping, eating, or working. | | | | | | | | | | | | |

### Supplemental Table 5. Age-standardized sociodemographic, health behavior, and clinical characteristics among college students, overall and by sex, National Health Interview Survey, 2022 (N=1,426)

|  | Total  n=1,426 (100%) | | | | Men  n=582 (40.8%) | | | | Women  n=844 (59.2%) | | | |
| --- | --- | --- | --- | --- | --- | --- | --- | --- | --- | --- | --- | --- |
|  |  | Life satisfaction ^a^ | | Chi-square or t-test  p-value |  | Life satisfaction ^a^ | | Chi-square or t-test  p-value |  | Life satisfaction ^a^ | | Chi-square or t-test  p-value |
|  | All n=1,426 (100%) | Yes n=1,385 (97.1%) | No n=41 (2.9%) |  | All n=582 (100%) | Yes n=561 (96.4%) | No n=21 (3.6%) |  | All n=844 (100%) | Yes n=824 (97.6%) | No n=20 (2.4%) |  |
| **Sociodemographic Characteristics** |  | | | |  | | | |  | | | |
| Age (years), mean (SE) | 27.7 (0.3) | 27.7 (0.3) | 26.9 (1.3) | 0.566 | 26.4 (0.4) | 26.5 (0.4) | 25.6 (1.6) | 0.589 | 28.6 (0.4) | 28.6 (0.4) | 28.8 (2.1) | 0.928 |
| Age group |  |  |  | 0.580 |  |  |  | 0.573 |  |  |  | 0.661 |
| <23 years | 45.8 | 46.0 | 40.5 |  | 51.4 | 51.7 | 44.3 |  | 41.4 | 41.5 | 35.1 |  |
| ≥23 years | 54.2 | 54.0 | 59.5 |  | 48.6 | 48.3 | 55.7 |  | 58.6 | 58.5 | 64.9 |  |
| Race and ethnicity |  |  |  | 0.390 |  |  |  | 0.514 |  |  |  | 0.557 |
| Hispanic/Latine | 14.8 | 14.7 | 20.8 |  | 11.0 | 10.9 | 16.4 |  | 16.7 | 16.7 | 22.3 |  |
| NH-Black/African American | 13.2 | 13.3 | 10.1 |  | 12.2 | 12.2 | 12.2 |  | 14.0 | 14.1 | 6.9 |  |
| NH-Other | 10.6 | 10.8 | 3.5 |  | 7.3 | 7.5 | 1.8 |  | 12.6 | 12.6 | 6.1 |  |
| NH-White | 61.4 | 61.2 | 65.6 |  | 69.5 | 69.5 | 69.5 |  | 56.7 | 56.5 | 64.7 |  |
| Student status |  |  |  | 0.273 |  |  |  | 0.135 |  |  |  | 0.902 |
| Undergraduate students | 58.5 | 58.5 | 65.5 |  | 55.8 | 55.6 | 74.2 |  | 59.5 | 59.6 | 56.8 |  |
| Graduate students | 41.5 | 41.5 | 34.5 |  | 44.2 | 44.4 | 25.8 |  | 40.5 | 40.4 | 43.2 |  |
| Employment status |  |  |  | 0.176 |  |  |  | 0.162 |  |  |  | 0.455 |
| Employed | 59.5 | 60.0 | 44.7 |  | 59.4 | 60.1 | 33.7 |  | 59.7 | 60.1 | 49.8 |  |
| Not employed | 40.5 | 40.0 | 55.3 |  | 40.6 | 39.9 | 66.3 |  | 40.3 | 39.9 | 50.2 |  |
| Ratio of family income to poverty threshold |  |  |  | 0.001 |  |  |  | 0.003 |  |  |  | <0.001 |
| <1.00 | 7.8 | 7.6 | 22.6 |  | 5.1 | 5.2 | 4.3 |  | 9.3 | 8.8 | 41.4 |  |
| 1.00-1.99 | 15.1 | 14.9 | 35.7 |  | 13.0 | 12.4 | 48.7 |  | 16.8 | 16.6 | 31.5 |  |
| 2.00-2.99 | 16.0 | 15.9 | 15.5 |  | 10.0 | 9.7 | 21.2 |  | 19.3 | 19.5 | 6.8 |  |
| ≥3.00 | 61.1 | 61.6 | 26.2 |  | 71.9 | 72.7 | 25.8 |  | 54.5 | 55.0 | 20.3 |  |
| Marital status |  |  |  | 0.002 |  |  |  | 0.029 |  |  |  | 0.161 |
| Divorced/Widowed/Separated/Married-spouse absent | 18.7 | 18.5 | 21.6 |  | 9.7 | 9.5 | 21.0 |  | 23.6 | 23.5 | 23.0 |  |
| Single/Never married | 36.9 | 36.5 | 66.0 |  | 40.9 | 40.5 | 71.2 |  | 34.2 | 34.0 | 57.6 |  |
| Married-spouse present/Living with partner | 44.4 | 44.9 | 12.4 |  | 49.4 | 50.0 | 7.8 |  | 42.2 | 42.6 | 19.4 |  |
| Region of residence |  |  |  | 0.715 |  |  |  | 0.179 |  |  |  | 0.678 |
| Northeast | 16.2 | 16.1 | 21.2 |  | 13.8 | 13.4 | 36.0 |  | 17.2 | 17.3 | 12.5 |  |
| Midwest | 22.0 | 22.0 | 24.2 |  | 16.5 | 16.7 | 7.5 |  | 24.9 | 24.7 | 35.3 |  |
| South | 34.7 | 35.0 | 24.0 |  | 40.3 | 40.7 | 22.3 |  | 32.4 | 32.7 | 20.0 |  |
| West | 27.1 | 26.9 | 30.6 |  | 29.5 | 29.3 | 34.2 |  | 25.4 | 25.3 | 32.1 |  |
| **Health Behaviors** |  | | | |  | | | |  | | | |
| Smoking status |  |  |  | 0.184 |  |  |  | 0.049 |  |  |  | 0.227 |
| Never/quit>12 months prior | 95.0 | 95.2 | 85.9 |  | 96.5 | 96.5 | 94.7 |  | 94.4 | 94.6 | 81.5 |  |
| Former/quit≤12 months ago | 0.8 | 0.8 | 2.3 |  | 0.8 | 0.7 | 3.9 |  | 0.8 | 0.8 | 0.0 |  |
| Current | 4.2 | 4.1 | 11.8 |  | 2.7 | 2.8 | 1.4 |  | 4.8 | 4.7 | 18.5 |  |
| Alcohol consumption |  |  |  | 0.445 |  |  |  | 0.582 |  |  |  | 0.181 |
| Current (≥1 drink past year) | 67.0 | 66.8 | 76.3 |  | 72.3 | 72.3 | 67.7 |  | 63.9 | 63.7 | 83.7 |  |
| Former (no drinks past year) | 17.2 | 17.2 | 10.3 |  | 11.5 | 11.6 | 4.7 |  | 20.5 | 20.5 | 12.3 |  |
| Lifetime abstinence (<12 drinks in life) | 15.8 | 15.9 | 13.4 |  | 16.1 | 16.0 | 27.5 |  | 15.6 | 15.8 | 4.0 |  |
| Leisure-time physical activity |  |  |  | 0.040 |  |  |  | 0.458 |  |  |  | 0.017 |
| Inactive | 18.0 | 17.6 | 41.0 |  | 12.5 | 12.2 | 28.5 |  | 21.6 | 21.1 | 53.1 |  |
| Insufficiently active | 25.2 | 25.2 | 24.2 |  | 17.5 | 17.3 | 27.6 |  | 30.0 | 30.0 | 26.1 |  |
| Sufficiently active | 56.8 | 57.2 | 34.8 |  | 70.1 | 70.5 | 43.9 |  | 48.4 | 48.9 | 20.8 |  |
| Usual sleep duration |  |  |  | 0.009 |  |  |  | 0.017 |  |  |  | 0.350 |
| Short (<7 hours) | 29.3 | 28.9 | 47.1 |  | 31.4 | 30.6 | 61.8 |  | 27.9 | 27.8 | 36.9 |  |
| Recommended (≥7 hours) | 70.7 | 71.1 | 52.9 |  | 68.6 | 69.4 | 38.2 |  | 72.1 | 72.2 | 63.1 |  |
| Few insomnia (yes) ^b^ | 76.6 | 77.3 | 37.4 | <0.001 | 76.7 | 77.9 | 34.7 | 0.010 | 76.2 | 76.7 | 39.5 | 0.005 |
| Restorative sleep (yes) ^c^ | 61.0 | 61.9 | 8.0 | <0.001 | 62.5 | 63.7 | 11.9 | <0.001 | 59.6 | 60.4 | 2.0 | <0.001 |
| **Clinical Characteristics** |  | | | |  | | | |  | | | |
| Ever had depression (yes) ^d^ | 23.2 | 22.4 | 68.5 | <0.001 | 20.0 | 19.3 | 55.3 | <0.001 | 25.8 | 24.8 | 88.7 | <0.001 |
| Body mass index category |  |  |  | 0.015 |  |  |  | 0.892 |  |  |  | <0.001 |
| Underweight (<18.5 kg/m^2^) | 1.5 | 1.4 | 9.6 |  | 1.4 | 1.5 | 0.0 |  | 1.5 | 1.3 | 19.0 |  |
| Recommended (18.5-<25 kg/m^2^) | 36.8 | 37.0 | 24.7 |  | 27.3 | 27.3 | 28.8 |  | 42.0 | 42.3 | 18.5 |  |
| Overweight (25-<30 kg/m^2^) | 32.7 | 32.6 | 39.5 |  | 39.9 | 39.8 | 40.2 |  | 28.3 | 28.3 | 34.9 |  |
| Obese (≥30 kg/m^2^) | 28.9 | 29.0 | 26.2 |  | 31.4 | 31.5 | 31.0 |  | 28.1 | 28.2 | 27.6 |  |
| General health status |  |  |  | <0.001 |  |  |  | 0.031 |  |  |  | <0.001 |
| Excellent/Very good/Good | 93.1 | 93.5 | 71.3 |  | 93.7 | 94.1 | 72.0 |  | 92.7 | 93.2 | 63.2 |  |
| Fair/poor | 6.9 | 6.5 | 28.7 |  | 6.3 | 5.9 | 28.0 |  | 7.3 | 6.8 | 36.8 |  |
| Abbreviations: SE (standard error), NH (non-Hispanic) | | | | | | | | | | | | |
| Note: All estimates are age-standardized to the US 2020 population, except for age and weighted for the survey's complex sampling design. Data are presented as column percentages or means and standard errors. Percentages may not sum to 100 due to rounding or missing data. College students are defined as having completed High School or more (EDUC) and currently attending school (SCHOOLNOW) | | | | | | | | | | | | |
| ^a^ Life satisfaction defined as a response of 'very dissatisfied' or 'dissatisfied' vs. 'satisfied' or 'very satisfied'. 'In general, how satisfied are you with your life? Are you very satisfied, satisfied, dissatisfied, or very dissatisfied?' | | | | | | | | | | | | |
| ^b^ Few insomnia defined as not having trouble falling and staying asleep 'most days' or 'every day' in past 30 days | | | | | | | | | | | | |
| ^c^ Restorative sleep defined as feeling rested upon waking 'most days' or 'every day' in past 30 days | | | | | | | | | | | | |
| ^d^ Depression was defined by the 2019 Field Representative's Manual defines depression (major depressive disorder or clinical depression) as a common but serious mood disorder. It causes severe symptoms that affect how you feel, think, and handle daily activities, such as sleeping, eating, or working. | | | | | | | | | | | | |

### Supplemental Table 6. Age-standardized sociodemographic, health behavior, and clinical characteristics among college students, overall and by student status, National Health Interview Survey, 2022 (N=1,426)

|  | Total  n=1,426 (100%) | | | | Undergraduate students  n=898 (63.0%) | | | | Graduate students  n=528 (37.0%) | | | |
| --- | --- | --- | --- | --- | --- | --- | --- | --- | --- | --- | --- | --- |
|  |  | Life satisfaction ^a^ | | Chi-square or t-test  p-value |  | Life satisfaction ^a^ | | Chi-square or t-test  p-value |  | Life satisfaction ^a^ | | Chi-square or t-test  p-value |
|  | All  n=1,426 (100%) | Yes n=1,385 (97.1%) | No  n=41 (2.9%) |  | All  n=898 (100%) | Yes  n=869 (96.8%) | No  n=29 (3.2%) |  | All  n=528 (100%) | Yes  n=516 (97.7%) | No  n=12 (2.3%) |  |
| **Sociodemographic Characteristics** |  | | | |  | | | |  | | | |
| Age (years), mean (SE) | 27.7 (0.3) | 27.7 (0.3) | 26.9 (1.3) | 0.566 | 25.2 (0.3) | 25.2 (0.3) | 25.1 (1.3) | 0.874 | 34.7 (0.6) | 34.7 (0.6) | 34.1 (3.3) | 0.980 |
| Age group |  |  |  | 0.580 |  |  |  | 0.399 |  |  |  | 0.378 |
| <23 years | 45.8 | 46.0 | 40.5 |  | 59.0 | 59.3 | 50.4 |  | 8.9 | 9.1 | 0.0 |  |
| ≥23 years | 54.2 | 54.0 | 59.5 |  | 41.0 | 40.7 | 49.6 |  | 91.1 | 90.9 | 100.0 |  |
| Sex |  |  |  | 0.136 |  |  |  | 0.055 |  |  |  | 0.741 |
| Men | 38.6 | 38.4 | 50.2 |  | 35.6 | 35.3 | 57.2 |  | 38.2 | 38.2 | 33.6 |  |
| Women | 61.4 | 61.6 | 49.8 |  | 64.4 | 64.7 | 42.8 |  | 61.8 | 61.8 | 66.4 |  |
| Race and ethnicity |  |  |  | 0.390 |  |  |  | 0.335 |  |  |  | 0.390 |
| Hispanic/Latine | 14.8 | 14.7 | 20.8 |  | 18.6 | 18.7 | 16.6 |  | 7.9 | 7.7 | 19.0 |  |
| NH-Black/African American | 13.2 | 13.3 | 10.1 |  | 13.3 | 13.3 | 11.2 |  | 13.4 | 13.6 | 4.7 |  |
| NH-Other | 10.6 | 10.8 | 3.5 |  | 16.0 | 16.2 | 2.3 |  | 9.2 | 9.2 | 9.4 |  |
| NH-White | 61.4 | 61.2 | 65.6 |  | 52.0 | 51.8 | 69.9 |  | 69.5 | 69.5 | 66.8 |  |
| Employment status |  |  |  | 0.176 |  |  |  | 0.204 |  |  |  | 0.226 |
| Employed | 59.5 | 60.0 | 44.7 |  | 57.0 | 57.5 | 33.9 |  | 64.9 | 65.4 | 47.2 |  |
| Not employed | 40.5 | 40.0 | 55.3 |  | 43.0 | 42.5 | 66.1 |  | 35.1 | 34.6 | 52.8 |  |
| Ratio of family income to poverty threshold |  |  |  | 0.001 |  |  |  | 0.039 |  |  |  | <0.001 |
| <1.00 | 7.8 | 7.6 | 22.6 |  | 9.8 | 9.4 | 35.2 |  | 5.4 | 5.3 | 16.3 |  |
| 1.00-1.99 | 15.1 | 14.9 | 35.7 |  | 21.2 | 21.2 | 27.8 |  | 8.6 | 7.9 | 46.7 |  |
| 2.00-2.99 | 16.0 | 15.9 | 15.5 |  | 21.0 | 21.0 | 15.5 |  | 10.3 | 10.2 | 15.5 |  |
| ≥3.00 | 61.1 | 61.6 | 26.2 |  | 47.9 | 48.3 | 21.5 |  | 75.7 | 76.6 | 21.5 |  |
| Marital status |  |  |  | 0.002 |  |  |  | <0.001 |  |  |  | 0.004 |
| Divorced/Widowed/Separated/Married-spouse absent | 18.7 | 18.5 | 21.6 |  | 20.6 | 20.8 | 2.0 |  | 17.9 | 17.5 | 37.5 |  |
| Single/Never married | 36.9 | 36.5 | 66.0 |  | 40.9 | 40.5 | 84.4 |  | 25.3 | 24.8 | 55.6 |  |
| Married-spouse present/Living with partner | 44.4 | 44.9 | 12.4 |  | 38.5 | 38.8 | 13.5 |  | 56.8 | 57.7 | 6.9 |  |
| Region of residence |  |  |  | 0.715 |  |  |  | 0.154 |  |  |  | 0.162 |
| Northeast | 16.2 | 16.1 | 21.2 |  | 13.4 | 13.2 | 34.2 |  | 17.7 | 17.9 | 5.0 |  |
| Midwest | 22.0 | 22.0 | 24.2 |  | 24.6 | 24.4 | 32.6 |  | 22.6 | 22.7 | 16.1 |  |
| South | 34.7 | 35.0 | 24.0 |  | 35.7 | 36.0 | 18.0 |  | 33.9 | 34.2 | 25.5 |  |
| West | 27.1 | 26.9 | 30.6 |  | 26.2 | 26.4 | 15.3 |  | 25.8 | 25.2 | 53.3 |  |
| **Health Behaviors** |  | | | |  | | | |  | | | |
| Smoking status |  |  |  | 0.184 |  |  |  | 0.223 |  |  |  | 0.648 |
| Never/quit>12 months prior | 95.0 | 95.2 | 85.9 |  | 92.2 | 92.4 | 72.7 |  | 97.6 | 97.6 | 94.9 |  |
| Former/quit≤12 months ago | 0.8 | 0.8 | 2.3 |  | 1.3 | 1.2 | 2.8 |  | 0.5 | 0.5 | 0.0 |  |
| Current | 4.2 | 4.1 | 11.8 |  | 6.5 | 6.3 | 24.5 |  | 1.9 | 1.8 | 5.1 |  |
| Alcohol consumption |  |  |  | 0.445 |  |  |  | 0.003 |  |  |  | 0.506 |
| Current (≥1 drink past year) | 67.0 | 66.8 | 76.3 |  | 56.8 | 56.4 | 88.9 |  | 78.9 | 79.1 | 71.7 |  |
| Former (no drinks past year) | 17.2 | 17.2 | 10.3 |  | 22.2 | 22.3 | 4.4 |  | 13.9 | 13.9 | 11.4 |  |
| Lifetime abstinence (<12 drinks in life) | 15.8 | 15.9 | 13.4 |  | 21.0 | 21.3 | 6.7 |  | 7.2 | 7.1 | 16.9 |  |
| Leisure-time physical activity |  |  |  | 0.040 |  |  |  | 0.114 |  |  |  | 0.009 |
| Inactive | 18.0 | 17.6 | 41.0 |  | 22.7 | 22.2 | 52.6 |  | 12.3 | 11.9 | 36.1 |  |
| Insufficiently active | 25.2 | 25.2 | 24.2 |  | 29.0 | 29.3 | 9.9 |  | 24.7 | 24.5 | 42.2 |  |
| Sufficiently active | 56.8 | 57.2 | 34.8 |  | 48.3 | 48.5 | 37.5 |  | 63.0 | 63.6 | 21.7 |  |
| Usual sleep duration |  |  |  | 0.009 |  |  |  | 0.050 |  |  |  | 0.550 |
| Short (<7 hours) | 29.3 | 28.9 | 47.1 |  | 29.1 | 28.6 | 43.1 |  | 29.5 | 29.4 | 38.0 |  |
| Recommended (≥7 hours) | 70.7 | 71.1 | 52.9 |  | 70.9 | 71.4 | 56.9 |  | 70.5 | 70.6 | 62.0 |  |
| Non-insomnia (yes) ^b^ | 76.6 | 77.3 | 37.4 | <0.001 | 78.3 | 79.2 | 36.4 | 0.053 | 76.6 | 77.1 | 46.0 | 0.009 |
| Restorative sleep (yes) ^c^ | 61.0 | 61.9 | 8.0 | <0.001 | 58.8 | 59.7 | 7.6 | <0.001 | 63.0 | 63.8 | 9.7 | <0.001 |
| **Clinical Characteristics** |  | | | |  | | | |  | | | |
| Ever had depression (yes) ^d^ | 23.2 | 22.4 | 68.5 | <0.001 | 23.7 | 23.0 | 66.4 | <0.001 | 22.6 | 21.6 | 78.3 | <0.001 |
| Body mass index category |  |  |  | 0.015 |  |  |  | 0.590 |  |  |  | 0.024 |
| Underweight (<18.5 kg/m^2^) | 1.5 | 1.4 | 9.6 |  | 1.8 | 1.7 | 5.0 |  | 0.8 | 0.7 | 9.2 |  |
| Recommended (18.5-<25 kg/m^2^) | 36.8 | 37.0 | 24.7 |  | 40.1 | 40.2 | 26.7 |  | 33.0 | 33.3 | 15.5 |  |
| Overweight (25-<30 kg/m^2^) | 32.7 | 32.6 | 39.5 |  | 29.8 | 29.7 | 37.0 |  | 37.2 | 37.0 | 49.8 |  |
| Obesity (≥30 kg/m^2^) | 28.9 | 29.0 | 26.2 |  | 28.4 | 28.4 | 31.3 |  | 28.9 | 29.0 | 25.5 |  |
| General health status |  |  |  | <0.001 |  |  |  | 0.304 |  |  |  | <0.001 |
| Excellent/Very good/Good | 93.1 | 93.5 | 71.3 |  | 90.5 | 90.6 | 86.5 |  | 95.6 | 96.4 | 49.1 |  |
| Fair/poor | 6.9 | 6.5 | 28.7 |  | 9.5 | 9.4 | 13.5 |  | 4.4 | 3.6 | 50.9 |  |
| Abbreviations: SE (standard error), NH (non-Hispanic) | | | | | | | | | | | | |
| Note: All estimates are age-standardized to the US 2020 population, except for age and weighted for the survey's complex sampling design. Data are presented as column percentages or means and standard errors. Percentages may not sum to 100 due to rounding or missing data. College students are defined as having completed High School or more (EDUC) and currently attending school (SCHOOLNOW) | | | | | | | | | | | | |
| ^a^ Life satisfaction defined as a response of 'very dissatisfied' or 'dissatisfied' vs. 'satisfied' or 'very satisfied'. 'In general, how satisfied are you with your life? Are you very satisfied, satisfied, dissatisfied, or very dissatisfied?' | | | | | | | | | | | | |
| ^b^ Non-insomnia defined as not having trouble falling and staying asleep 'most days' or 'every day' in past 30 days | | | | | | | | | | | | |
| ^c^ Restorative sleep defined as feeling rested upon waking 'most days' or 'every day' in past 30 days | | | | | | | | | | | | |
| ^d^ Depression was defined by the 2019 Field Representative's Manual defines depression (major depressive disorder or clinical depression) as a common but serious mood disorder. It causes severe symptoms that affect how you feel, think, and handle daily activities, such as sleeping, eating, or working. | | | | | | | | | | | | |

### Supplemental Table 7. Prevalence of life satisfaction among college students by race and ethnicity and the student status, National Health Interview Survey, 2022, (N=1,426)

|  | Total | | | | Hispanic/Latine | | | | NH-Black/African American | | | | NH-Other^*^ | | | | NH-White | | | |
| --- | --- | --- | --- | --- | --- | --- | --- | --- | --- | --- | --- | --- | --- | --- | --- | --- | --- | --- | --- | --- |
|  | All n=1,426 (100%) | Undergraduate students n=898 (63.0%) | Graduate students n=528 (37.0%) | Chi-square or t-test p-value | All n=299 (100%) | < Undergraduate students n=239 (79.9%) | Graduate students n=60 (20.1%) | Chi-square or t-test p-value | All n=200 (100%) | Undergraduate students n=129 (64.5%) | Graduate students n=71 (35.5%) | Chi-square or t-test p-value | All n=200 (100%) | Undergraduate students n=115 (57.5%) | Graduate students n=85 (42.5%) | Chi-square or t-test p-value | All n=727 (100%) | Undergraduate students n=415 (57.1%) | Graduate students n=312 (42.9%) | Chi-square or t-test p-value |
| Life satisfaction ^a^ |  |  |  | 0.273 |  |  |  | 0.340 |  |  |  | 0.031 |  |  |  | 0.487 |  |  |  | 0.265 |
| Dissatisfied | 1.9 | 1.9 | 1.6 |  | 2.7 | 2.0 | 4.4 |  | 1.6 | 2.0 | 0.5 |  | 0.6 | 0.5 | 1.1 |  | 2.1 | 2.3 | 1.7 |  |
| Satisfied | 98.1 | 98.1 | 98.4 |  | 97.3 | 98.0 | 95.6 |  | 98.4 | 98.0 | 99.5 |  | 99.4 | 99.5 | 98.9 |  | 97.9 | 97.7 | 98.3 |  |
| Abbreviations: NH (non-Hispanic) | | | | | | | | | | | | | | | | | | | | |
| Note: All estimates are age-standardized to the US 2020 population, except for age and weighted for the survey's complex sampling design. Data are presented as column percentages or means and standard errors. Percentages may not sum to 100 due to rounding or missing data. College students are defined as having completed High School or more (EDUC) and currently attending school (SCHOOLNOW) | | | | | | | | | | | | | | | | | | | | |
| ^a^ Life satisfaction defined as a response of 'very dissatisfied' or 'dissatisfied' vs. 'satisfied' or 'very satisfied'. 'In general, how satisfied are you with your life? Are you very satisfied, satisfied, dissatisfied, or very dissatisfied?' | | | | | | | | | | | | | | | | | | | | |
| ^*^ NH-Other includes non-Hispanic, Asian, American Indian/Alaska native, Multiple and other races. | | | | | | | | | | | | | | | | | | | | |
